# Quantifying Asymptomatic Infection and Transmission of COVID-19 in New York City using Observed Cases, Serology and Testing Capacity

**DOI:** 10.1101/2020.10.16.20214049

**Authors:** Rahul Subramanian, Qixin He, Mercedes Pascual

## Abstract

The contributions of asymptomatic infections to herd immunity and community transmission are key to the resurgence and control of COVID-19, but are difficult to estimate using current models that ignore changes in testing capacity. Using a model that incorporates daily testing information fit to the case and serology data from New York City, we show that the proportion of symptomatic cases is low, ranging from 13% to 18%, and that the reproductive number may be larger than often assumed. Asymptomatic infections contribute substantially to herd immunity, and to community transmission together with pre-symptomatic ones. If asymptomatic infections transmit at similar rates than symptomatic ones, the overall reproductive number across all classes is larger than often assumed, with estimates ranging from 3.2 to 4.4. If they transmit poorly, then symptomatic cases have a larger reproductive number ranging from 3.9 to 8.1. Even in this regime, pre-symptomatic and asymptomatic cases together comprise at least 50% of the force of infection at the outbreak peak. We find no regimes in which all infection sub-populations have reproductive numbers lower than 3. These findings elucidate the uncertainty that current case and serology data cannot resolve, despite consideration of different model structures. They also emphasize how temporal data on testing can reduce and better define this uncertainty, as we move forward through longer surveillance and second epidemic waves. Complementary information is required to determine the transmissibility of asymptomatic cases, which we discuss. Regardless, current assumptions about the basic reproductive number of SARS-Cov-2 should be reconsidered.

**Significance Statement:** As health officials face another wave of COVID-19, they require estimates of the proportion of infected cases that develop symptoms, and the extent to which symptomatic and asymptomatic cases contribute to community transmission. Recent asymptomatic testing guidelines are ambiguous. Using an epidemiological model that includes testing capacity, we show that most infections are asymptomatic but contribute substantially to community transmission in the aggregate. Their individual transmissibility remains uncertain. If they transmit as well as symptomatic infections, the epidemic may spread at faster rates than current models often assume. If they do not, then each symptomatic case generates on average a higher number of secondary infections than typically assumed. Regardless, controlling transmission requires community-wide interventions informed by extensive, well-documented asymptomatic testing.

---

**S**ince the emergence of the novel coronavirus in December 2019(1), the COVID-19 pandemic has resulted in over 16 million cases and 600,000 deaths worldwide(2). Schools and universities in the United States are gradually re-opening amid concerns that a second wave of the epidemic may re-emerge in the fall and winter of 2020.

As they craft testing policies and intervention strategies to mitigate a second wave, public health officials need to better understand the role that symptomatic and asymptomatic individuals play in the community transmission of COVID-19 and in the development of herd immunity to the disease. However, fundamental epidemiological questions remain poorly understood, including what fraction of cases are symptomatic and how well asymptomatic cases can transmit relative to symptomatic ones. These questions are especially urgent given ambiguity in recent CDC guidelines regarding the testing of asymptomatic individuals(3).

Answering these questions can also provide further insight on the basic reproductive number of SARS-CoV-2, and how the virus would spread in a population in the absence of interventions. This number known as *R*_0_ is defined as the mean number of secondary cases arising from a primary case in the absence of immunity, and is estimated on the basis of a particular epidemiological model. Mathematical models for the population dynamics of COVID-19 incorporate different features such as asymptomatic and pre-symptomatic transmission, super-spreading, or heterogeneity in susceptibility. A considerable range of *R*_0_ estimates has been reported, ranging from at least 1.5(4) to 5.7(5) in Wuhan. A much narrower range between 2 and 3 is frequently cited in the popular press, or assumed when simulating models(6)or fitting these to data(7, 8). This assumption may be based on the dynamics of COVID-19 in regions that implemented interventions early(9–13). A more precise estimate of *R*_0_ from a city where substantial transmission was occurring prior to intervention, such as New York City, would provide a relevant baseline. Furthermore, if “super-spreading” by a small fraction of symptomatic infections fuels COVID-19 transmission, a precise estimate of the mean number of secondary cases arising from such an individual, may be just as valuable. A model that precisely estimates the fraction of symptomatic cases may help epidemiologists discern if either the overall or symptomatic reproductive numbers are higher than assumed.

The probability that a COVID-19 infection is symptomatic is difficult to estimate(14) and a wide range of values have been suggested (14–16). Estimates from cruise ship outbreaks(17), Wuhan evacuees(18), long term care facilities(19), and contact tracing of index cases(15) may not be representative of the general population. Increases in the testing capacity for COVID-19 over time(9, 20, 21) make population-level estimation of this probability difficult due to confounding with other parameters such as the reporting, hospitalization, and fatality rates. When the testing capacity is limited in the early stages of an outbreak, severe cases are more likely to be tested, which can bias estimates of the probability that an infection is symptomatic and the fatality rate. Changes in testing capacity over time also confound the definition itself of asymptomatic individuals in transmission models, when these are not differentiated from unreported cases. These changes can also bias the reported deaths attributed to COVID-19.

These challenges can be improved upon by explicitly incorporating changes in testing capacity into an epidemiological process model. While some early models of the COVID-19 outbreak in Wuhan attempted to take into account changes in testing capacity(21) or differences in reporting rate during periods of the epidemic(9), the limited information on these trends in Wuhan meant that they had to be estimated on a coarse temporal scale (2-3 week intervals) and had to be inferred along with other parameters in the model. In the United States, many states and municipalities such as New York City(22, 23) have published daily estimates of the number of total COVID-19 tests conducted, together with the number of positive COVID-19 tests. While these data are often used by public health officials to gauge the spread of the COVID-19 outbreak, they have yet to be incorporated explicitly into epidemiological models.

We present an epidemiological model that incorporates RT-PCR testing as an integral process informed by empirical levels. The explicit consideration of testing allows us to clearly define asymptomatic individuals as those that will never transition to displaying symptoms, and to differentiate them from those who have been unreported because they were not tested. We fit the model to PCR-confirmed COVID-19 cases in New York City, using publicly available data provided by the New York State Department of Public Health(23). The resulting model can clearly delineate symptomatic and asymptomatic infections independently from the reporting rate. We subsequently fit the model to estimates of herd immunity obtained from a recent serological study in New York City(24) to further constrain inference results.

Our model obtains a precise estimate for the symptomatic proportion of COVID-19 cases. We show that most COVID-19 infections are asymptomatic, and that these asymptomatic infections together with pre-symptomatic ones substantially drive community transmission, contributing 50% or more of the total force of infection. Furthermore, depending on the transmissibility of individual asymptomatic cases relative to symptomatic ones, either the overall reproductive number or the symptomatic reproductive number may be higher than typically assumed. Our results highlight the importance of testing and contact tracing of asymptomatic individuals, and of making these data publicly available as health officials prepare for and manage a second wave.

## Results

We present a stochastic epidemiological model (Fig. 1) that explicitly incorporates daily changes in testing capacity and the lag between sampling and testing (see Methods). The underlying model, referred to hereafter as the SEPIAR model (Fig. 1A) has a susceptible-exposed-infectious-recovered structure with compartments for both severe (hospitalized) and non-severe symptomatic infections as well as pre-symptomatic (P) and asymptomatic (A) infections. We also consider two nested simplified versions: one with no pre-symptomatic transmission (the SEIAR model, Fig. 1B); and one with no asymptomatic transmission (the SEPIR model, Fig. 1C). By varying specific parameters weighting the transmission rate of P and A relative to that of symptomatic individuals, we can continuously move across these two extreme structures. Daily reports of the number of tests conducted in New York City are fed in as a covariate in the testing sub-model (see SI Appendix). The model takes into account CDC priorities in sampling and testing: all hospitalized cases are sampled and eventually tested, while non-severe symptomatic individuals are sampled and tested only if excess capacity is available at the time of sampling. We also incorporate the re-testing of hospitalized individuals as they leave the hospital. This model is fit to observed cases in New York City from March 1,2020 to June 1, 2020 and serological estimates of herd immunity in New York City from March 8,2020 to April 19,2020 (see Methods and SI Appendix). We compare the full model with the two nested simplified versions. Although all three model structures are supported by the case data, the model with no asymptomatic transmission is not supported when these data are considered in conjunction with serology information (SI Appendix, Table S2).

**Fig. 1.**
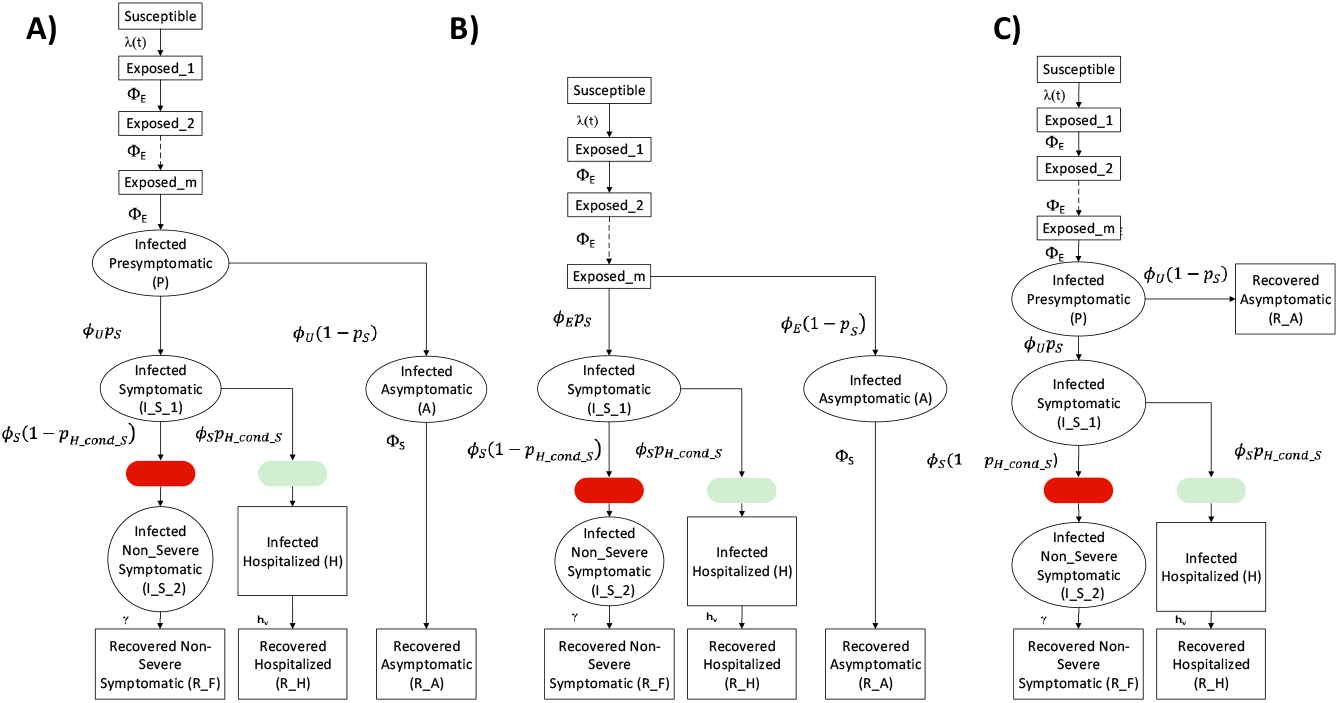
Model diagrams. (A) The full SEPIAR model used for inference. The model is an extension of an SEIR formulation that considers both pre-symptomatic transmission (from compartment *P*) and asymptomatic transmission (from compartment *A*). **B) When the strength of pre-symptomatic transmission** *b*_p_ **is set to 0, the SEPIAR model reduces to the SEIAR model**. Since we assume that *ϕ*_*U*_ = *ϕ*_*E*_, when *b*_*p*_ = 0 the infectious pre-symptomatic compartment behaves like an additional exposed compartment. **C) When the strength of asymptomatic transmission** *b*_a_ **is set to 0, the SEPIAR model reduces to the SEPIR model**. Individuals in the asymptomatic infectious compartment (*A*) make no contribution to the force of infection, so asymptomatic individuals essentially recover after leaving the pre-symptomatic period (*P*). In all three panels, circular/elliptical compartments contribute to the force of infection, while rectangular compartments do not. The green ellipse denotes the point at which severe/hospitalized COVID patients are sampled and enter the testing queue for severe cases, while the red ellipse denotes the corresponding entry point for the queue for non-severe symptomatic cases.

To evaluate the strength of transmission in asymptomatic cases relative to symptomatic cases, we construct a Monte Carlo profile using the f ull SEPIAR model (SI Appendix, Fig. S6). We isolate parameter combinations from the profile that are supported by the case and serology data, and examine the values of those combinations. Particular parameters of interest that we focus on include the proportion of cases that are symptomatic, *p*_*S*_, the ratio of the transmission rate of asymptomatic individuals to that of symptomatic individuals, *b*_a_, and the reproductive numbers. We use *R*_0_ to denote the symptomatic reproductive number (i.e. the mean number of secondary cases arising from each primary symptomatic case), and 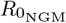 to denote the overall reproductive number for the model (i.e. the mean number of cases arising from a primary infection, where the average considers all types of infections).

The proportion of COVID-19 cases that are symptomatic is well identified, w ith a c onfidence interval ranging fr om 12.9% to 17.4% (Figure 2). Although a wide range of parameter combinations for the proportion of symptomatic infection are supported by the case data on its own, a much narrower estimate is obtained when the case and serology data are considered together (Fig. 2A, B). Within this range, estimates of herd immunity are consistent with the dynamics of observed serology (Fig. 2C), in particular the rapid rise in seroprevalence over March and April 2020.

**Fig. 2.**
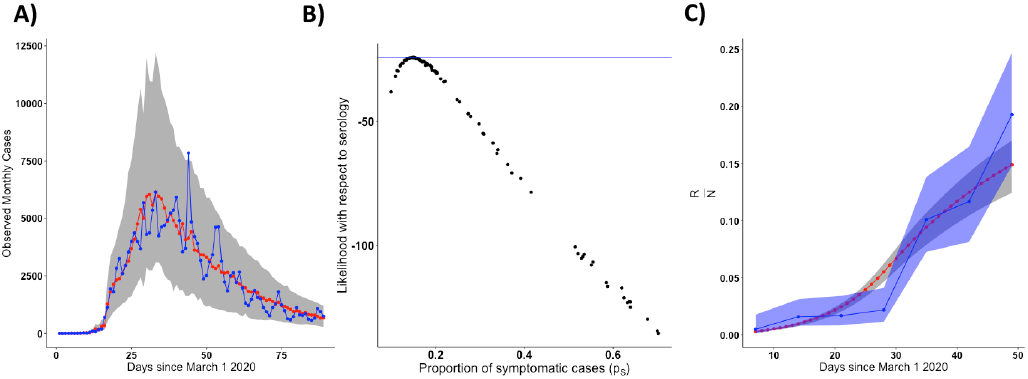
The probability of symptomatic infection. (A) Simulated vs. observed cases from the profile of the asymptomatic transmission strength (*b*_a_) using the SEPIAR model. The red line is the median from 100 simulations using the Maximum-Likelihood Estimates (MLE), while the grey shaded region denotes the 2.5-97.5% quantiles across 100 simulations from all parameter combinations within 2 log-likelihood units of the profile MLE. Likelihoods here are with respect to case data. The observed daily case counts are denoted by the blue line. **B) Model Likelihood as a function of the proportion of cases that are symptomatic (***p*_*S*_ **) for each parameter combination from panel A**. The y-axis shows the likelihood for that parameter combination with respect to serology data. All parameter combinations above the blue line have likelihoods within 2-log-likelihood units of the MLE (defined with respect to serology). This corresponds to a range of values for *p*_*S*_ of approximately 13-18%. **C) Comparison of observed vs. simulated estimates of herd immunity in the population from parameter combinations supported by both case and antibody data (all points above the blue line in panel B)**. The red line denotes the median value of herd immunity (the proportion of the population that has recovered 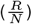 at that point in time in 100 simulations from the MLE parameter combination. The grey shaded region denotes the 2.5-97.5% quantiles for these simulations from all parameter combinations within 2-log-likelihood units of the MLE with respect to serology (all parameter combinations above the blue line in panel B). The blue line denotes estimates of herd immunity from a recent serological survey in New York City(24). The blue shading denotes 95% confidence intervals for those serology estimates using the methods of (24).

The overall reproductive number or symptomatic reproductive number may be larger than is often assumed. From our profile of the relative asymptomatic transmission r ate *b*_a_, we identify two main regimes of transmission that are supported by both the case and serology data (Fig. 3), in which either 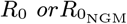 is higher than the 2-3 range often assumed for COVID-19. Notably, we find no parameter combinations in which both reproductive numbers are below 3 and fall within this range.

**Fig. 3.**
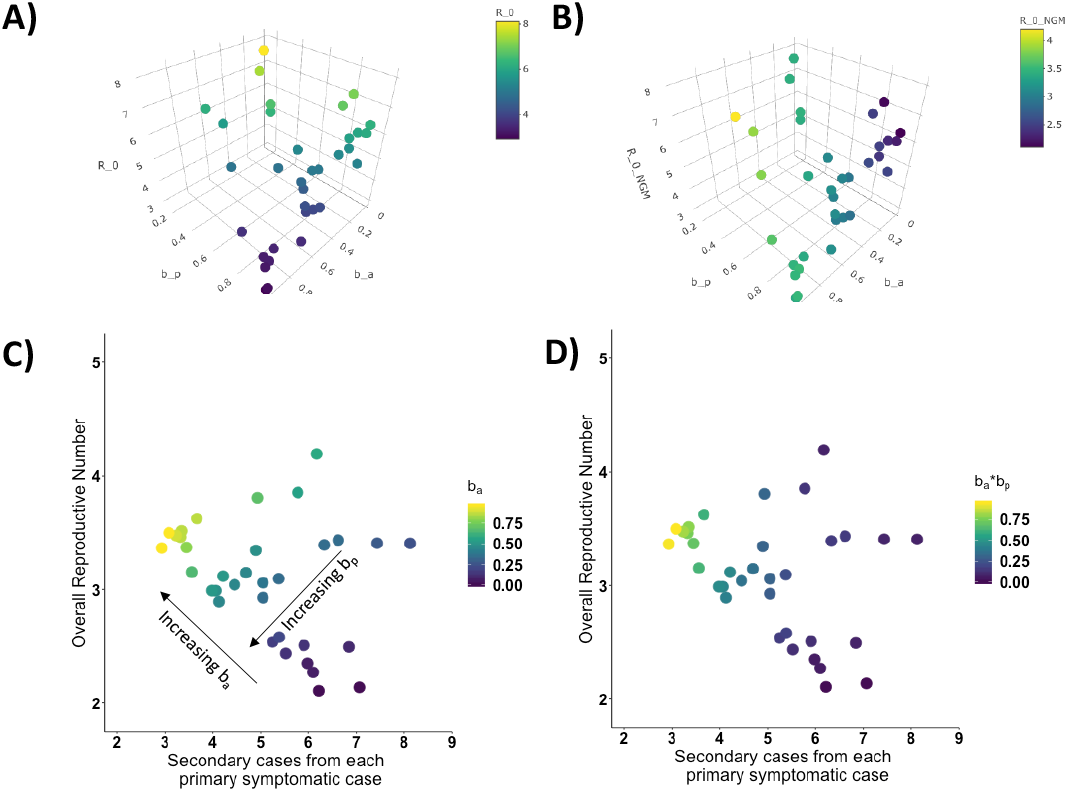
Surface plots of the reproductive number of symptomatic individuals (*R*_0_) (A) and the overall reproductive number 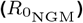, as a function of the relative strength of pre-symptomatic transmission (*b*_p_) and the relative strength of asymptomatic transmission (*b*_a_). Each point represents one parameter combination within 2 log-likelihood units of the MLE (with respect to serology) from the *b*_a_ profile. **C) Plot of the overall reproductive number vs the reproductive number in symptomatic individuals for the same points colored by** *b*_a_. The black arrows show the direction of increasing strength of asymptomatic transmission (*b*_a_) and pre-symptomatic transmission (*b*_p_). For this same plot except colored by the strength of pre-symptomatic transmission (*b*_p_), see SI Appendix Fig. S7. **D) The same plot except colored by the product of the strength of pre-symptomatic transmission (***b*_p_ **) multiplied by the strength of asymptomatic transmission (***b*_a_ ). For ease of plotting, we exclude two parameter combinations which had a very low relative rates of pre-symptomatic transmission (i.e. *b*_*p*_ was lower than 0.020). The two outlier combinations had high reproductive numbers (*R*_0_ = 17.77, 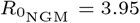 and *R*_0_ = 4.97, 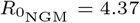). These outliers are included in the SI Appendix Fig. S8.

In the first regime, asymptomatic individuals transmit at almost the same rate as symptomatic individuals. That is, *b*_a_ is large, even close to 1 in some parameter combinations. The overall reproductive number takes on values between 3.2 and 4.4, and asymptomatic cases substantially contribute to the overall force of infection (Fig. 4).

**Fig. 4.**
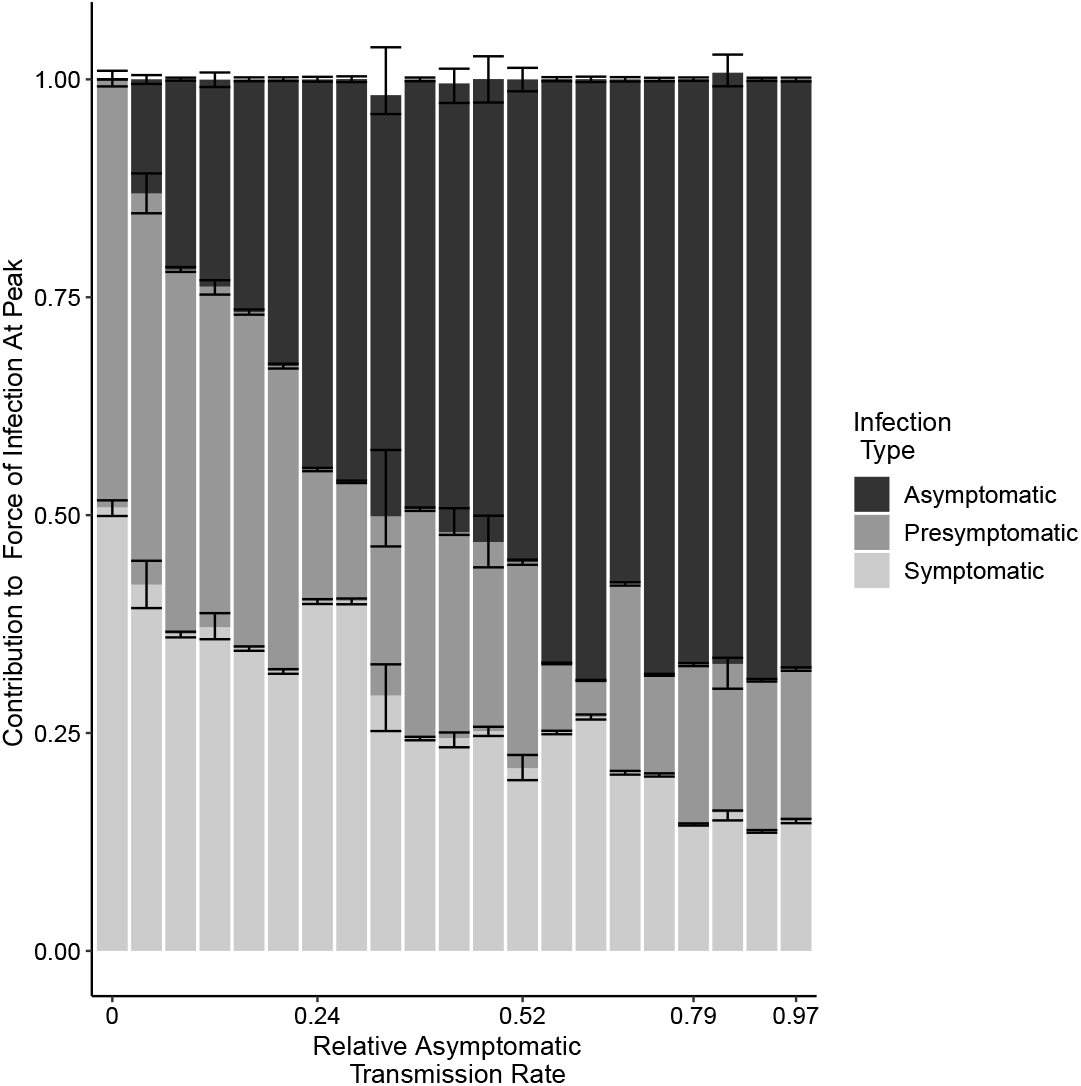
The contribution to the force of infection at the peak of the outbreak on April 14, 2020 from symptomatic, asymptomatic, and pre-symptomatic infections under different relative asymptomatic transmission rates *b*_a_. For each parameter combination from the fitted SEPIAR model supported by case and serology data (corresponding to the points in Figure 3), we simulate 100 trajectories and calculate the proportion of the overall force of infection on April 14,2020 that is due to asymptomatic, symptomatic, and pre-symptomatic infections. We pool trajectories from all parameter combinations that have the same value of *b*_*a*_, and calculate the median, 2.5%, and 97.5% quantiles for each infection class and value of *b*_*a*_. The colored bars represent for each infection class, the median proportion of its contribution to the force of infection (and hence may not sum exactly to 1). The error bars represent the corresponding 2.5%, and 97.5% quantiles. Versions of this plot calculated respectively 4 weeks before, and 4 weeks after, the peak can be found in the SI Appendix Fig. S10. We excluded two outlier parameter combinations that had extremely low relative rates of pre-symptomatic transmission (i.e. where *b*_p_ was less than 0.02).

In the second regime, asymptomatic individuals transmit at very low rates relative to symptomatic individuals, with estimates of *b*_a_ close to zero or in some parameter combinations even equal to zero. Concomitantly, the symptomatic reproductive number is much higher than frequently assumed, taking on values between 3.9 and 8.1. Nevertheless, even in this regime pre-sympomatic and asymptomatic infections together contribute at least 50% of the overall force of infection at the peak of the outbreak.

In both regimes, pre-symptomatic individuals transmit at almost the same rate as symptomatic individuals, with estimates of *b*_p_ close to 1, also making a substantial contribution to the overall force of infection (Fig. 4).

We also observe a third regime in which both reproductive numbers are higher than assumed, but in this regime pre-symptomatic individuals transmit at a very low rate, with *b*_p_ close to 0. Several combinations in this regime can be observed in the top right corner of Fig. 3 (C,D) and in Fig. S8. This is also the regime obtained in Fig. S7 if one uses the SEIAR model, which assumes that pre-symptomatic individuals do not transmit (i.e. *b*_p_ is fixed at 0). Given previous evidence of pre-symptomatic transmission of COVID-19(25, 26), we focus on the two regimes which incorporate substantial pre-symptomatic transmission.

In line with previous studies(27), we estimate a large value for the initial number of infected and incubating individuals with COVID-19 in New York City at the start of the simulation on March 1st. Parameter combinations that were supported by the case and serology data ranged from 9,000-18,000 initial infected individuals and 44,000-72,000 exposed individuals. A key question to consider when evaluating the plausibility of this magnitude of undetected infections is whether it is consistent with no signal of an anomalous number of hospitalizations. In other words, would this large rise in early infections result in a corresponding rise in COVID-hospitalizations that may not have been detected as COVID-related? We examine this question by comparing simulated daily hospitalizations from our fitted model with observed COVID-19 daily hospitalizations in New York City, as well as with syndrome surveillance reports of respiratory illness from emergency departments in New York City hospitals (SI Appendix, Fig. S5), which we can use as an indicator for a rise in undetected hospitalizations. We show that a scenario with a large number of initial infections on March 1st is indeed consistent with the time at which observed COVID-19 hospitalizations peak, providing further support for this contention. We also find t hat the imposition of social distancing on March 17th and the stay at home order on March 22nd in New York City resulted in a substantial decrease in the initial transmission rate. Parameter estimates for the ratio of the post-intervention transmission rate to the pre-intervention transmission rate (*b*_q_) ranged from 0.134 to 0.240, corresponding to a 75.98%-86.62% reduction in the strength of transmission after the intervention.

## Discussion

With a transmission model that incorporates daily changes in testing capacity, we estimate that the probability that an exposed individual develops symptoms is low. Since asymptomatic infections represent a large fraction of the infected population, they contribute substantially to community transmission in the aggregate together with pre-symptomatic cases, even when they individually transmit at a low per-capita rate. They also contribute substantially to building herd immunity.

We use testing information to estimate the probability that a new case will become symptomatic without the biases present in cruise-ship(17) and traveler studies(18), or the parameter confounding present in city-wide models. Early cruise-ship and evacuee studies found that most COVID-19 cases were symptomatic. However, given the small number of total infections(18, 28), evacuee studies may over-estimate the fraction of symptomatic cases if infections in observed severe cases(29) last longer(30) than in asymptomatic ones. Cruise-ship studies may likewise over-estimate this parameter if asymptomatic cases, which were tested later than symptomatic cases(17), recover prior to testing. City-wide models, which avoid these biases, indicate that most COVID-19 cases are undetected (9). They confound however the fraction of symptomatic cases with the reporting or hospitalization rate, as they neglect daily testing changes, and cannot distinguish between asymptomatic and undetected cases. The alternative approach of fitting the models to death data i s not necessarily exempt from biases in parameter estimates, due to changes in hospital capacity over time(31, 32), co-morbidities in host populations(33, 34), and the long delay between the onset of infection and death(35). Furthermore, the under-reporting of cases can also bias the assumed case fatality rate(32). Our approach resolves these issues by incorporating daily testing capacity as part of the model when estimating parameters from serology and case data. Models without explicit consideration of this capacity have difficulty estimating the proportion of cases that are symptomatic from these data (36), suggesting that including testing is crucial.

If asymptomatic individuals transmit at a high rate, then the overall reproductive number pre-intervention in New York City is larger than the 2-3 range often assumed in models(6–8) and media reports (11, 37–39) based on early estimates from Wuhan (4, 40, 41). Furthermore, we find no supported parameter combinations in which both the overall and symptomatic reproductive numbers fall within this range. Early Wuhan models may under-estimate *R*_0_ by ignoring pre-symptomatic transmission and making restrictive assumptions, including that COVID-19 has the same incubation period and serial interval as SARS-CoV (4, 40, 41), or that most cases are symptomatic(42). Early Wuhan case data may be insufficient to precisely estimate *R*_0_ without making these assumptions (43–45). Thus, models and intervention strategies should consider that the overall *R*_0_ may be higher than 3 in certain locations (5, 46).

If asymptomatic individuals are unlikely to transmit and do so with low probability, then the small fraction of cases that are symptomatic are transmitting at a high rate, in line with recently reported “super-spreading” events(47, 48). Super-spreading events are instances in which a single infected individual infects a large number of people. These events can be hard to measure on a population level in the absence of detailed transmission data. In classic super-spreading dynamics, most primary cases do not result in many secondary cases, while a subset of primary cases result in a large number of secondary cases(8, 49, 50). This heterogeneity in the reproductive number is indeed what we observe when asymptomatic individuals transmit poorly. Our model is admittedly a coarse description of this heterogeneity, since it incorporates only two different classes of infections, symptomatic or asymptomatic. Future models can build upon this framework with additional classes for age, socio-economic status, location or susceptibility(51) using fine-scale case data. However, our results also indicate that even when the symptomatic reproductive number is large, pre-symptomatic and asymptomatic infections contribute together to at least 50% of the overall force of infection.

It follows that community-wide interventions that account for non-symptomatic cases should be crucial for mitigating outbreaks. If asymptomatic cases transmit poorly, then concurrent additional interventions targeting super-spreading symptomatic infections may help reduce community transmission.

Resolving the non-identifiability of the efficacy of asymptomatic transmission (*b*_*a*_), would require extensive community testing and contact tracing of asymptomatic cases. Community testing on its own can provide an estimate of the total proportion of cases that are asymptomatic, but it may not provide insight on whether those asymptomatic individuals can transmit and how well they can transmit. Symptomatic and asymptomatic individuals have similar viral loads(52), but a high viral load does not necessarily imply high transmissibility. One limitation of early contact tracing studies is that estimates of transmissibility may over-sample symptomatic index cases and contacts, particularly during the early phase of an epidemic(15, 53). In certain studies, only symptomatic contacts are further investigated. Ideally, one would use frequent systematic community testing for studies identifying both symptomatic and asymptomatic potential index cases for further contact tracing and testing of all contacts regardless of symptoms. Furthermore, fixing the probability that an infection becomes symptomatic based on the results of serology-informed models such as ours, could increase the precision with which contact tracing studies can estimate the strength of asymptomatic transmission. Colleges that are currently reopening may be ideal test locations for this kind of combined approach, which may also help detect super-spreading events.

While it cannot capture all testing intricacies, our framework illustrates how transmission models can incorporate daily changes in testing capacity and identify parameters that were previously difficult to estimate such as the probability that an infection will become symptomatic. While we do not explicitly denote differences between labs, hospitals, or diagnostic tests, we account for this variation by including additional measurement noise after simulating the RT-PCR testing process. We also consider how sampling individuals without COVID-19 may deplete the daily testing capacity. In particular, hospitalized individuals with non-COVID-19 related severe respiratory disease may have a higher priority for testing than non-severe COVID-19 cases. Our model uses syndrome surveillance reports(54–57) of respiratory illness from New York City hospitals in previous years, along with weekly influenza cases, to estimate the number of non-COVID-19 severe respiratory cases that were tested. This framework could be used in conjunction with other epidemiological models, and extended to other municipalities or countries with location-specific testing priorities, re-testing procedures, or diagnostic tests. It could also be used to examine how altering testing strategies such as switching from symptom-based testing to community testing may improve transmission parameter inference and efficacy of control efforts. This may be an important consideration for countries that have limited testing capacity but are still in the midst of the first pandemic wave, such as India.

Future studies can investigate the impact of including a testing sub-model on parameter estimation and the level of detail required in such a sub-model. For example, one could compare the results of parameter estimation from fitting a given epidemiological model with a queue-based testing model to those that assume a fixed reporting rate and a delay in the reporting of cases. We expect the former to exhibit more uncertainty when informed by surveillance data from the beginning of the pandemic when little testing capacity is available, but to reduce this uncertainty as the time series is extended and this capacity changes. Models that assume a fixed reporting rate may under-estimate the range in uncertainty of epidemiological parameters that are heavily informed by the early part of the time series, and may even under-estimate the values of the parameters themselves. Models with a queue-based testing sub-model may obtain more precise estimates of parameters that impact the end of the outbreak, such as those related to the depletion of susceptible individuals, acquisition of immunity, or in our model, the impact of social distancing and stay-at-home orders on overall transmission. Even if including some form of testing model that takes into account changes in capacity is key to obtaining more precise parameter estimates, simpler versions of our implementation may be sufficient. For example, the more generalizable components such as the testing of hospitalized individuals may be more important than taking into account their re-sampling as they leave the hospital. Simplifying the testing model based on model selection analyses can facilitate wider adaption of the testing framework to other cities, countries, or time periods.

Our finding that m any individuals were already infected by March 1st is consistent with earlier estimates that community transmission began in February in NYC(27, 55, 58). Previous studies could not explain however why no substantial increase in COVID-19-like illness was observed prior to February 28th in syndrome surveillance data(55). Our simulations show that the lag between infection onset and hospitalization can explain this discrepancy. Even when initialized with many infected cases on March 1st, simulated hospitalizations do not rise until several weeks later concurrent with observed COVID-19 hospitalizations (SI Appendix Fig. S5). Most likely, the estimated initial conditions suggest multiple parallel foci of initiation of the epidemic with multiple importations of infections. Another suggested possibility is a dosage-dependence effect, wherein the severity of an individual’s infection depends on the size of the virus population that the person becomes infected with during one or more transmission events, and hence on the overall viral load of COVID-19 in the community. In this scenario, early COVID-19 cases in February and early March would be less severe. This would be consistent with the syndrome surveillance data, where we see a rise in early March of respiratory infection reports in the emergency departments of hospitals, but do not yet see a rise in COVID-19 hospitalizations. This phenomenon might also explain why our model slightly under-estimates the peak in daily hospitalizations, even though it correctly identifies the t ime and shape of that peak.

In conclusion, explicit consideration of changes in testing capacity allows us to infer with certainty from case and serology data that most new COVID-19 cases do not become symptomatic. We also inferred that the overall or symptomatic reproductive number may be larger than often assumed depending on how well asymptomatic cases can transmit. Despite this uncertainty, the strong consistent contribution to community transmission from cases without symptoms observed across scenarios supported by the data, should be considered when formulating public health intervention strategies. Making available detailed information on testing policy and data on testing capacity over time will strengthen the ability of epidemiological models to learn from the past and inform us about the future.

## Materials and Methods

We examine three different model structures that have been used to characterize COVID-19 dynamics in previous studies (Fig 1). All models are modified versions of the traditional susceptible-exposed-infected-recovered (SEIR) model (59). The first model, the SEPIR model(17, 60), is the most standard extension in which transitions are between a linear chain of compartments. Its formulation adds a compartment P for pre-symptomatic transmission. The second one, the SEIAR model (7, 9), differs conceptually in that it includes asymptomatic individuals rather than pre-symptomatic ones, and defines them as distinct, in the sense that they will never transition to exhibiting symptoms. This definition implicitly recognizes that there are essentially two classes of individuals in terms of susceptibility to disease and symptoms. The third structure for the SEPIAR model(6, 26) is a combination of the first two and includes them as nested, particular, cases.

All three models include a chain of *m* exposed classes to incorporate the total time between the onset of infection and the onset of symptoms as gamma distributed (with mean 5.5. days and standard deviation 2.25 days) (61). Symptomatic individuals are subdivided into two sequential classes 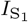 and 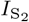 for practical purposes, to follow their numbers before and after some of them transition to hospitalization. Individuals spend an average of 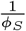 days in 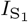 and 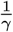 days in 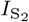.

The parameter *R*_0_ represents the reproductive number experienced by symptomatic individuals. We define a baseline pre-intervention transmission rate in symptomatic individuals *β*_0_ by dividing *R*_0_ by the average total time that non-severe cases transmit with symptoms. We also define a post-intervention transmission rate *β*_1_, which is equal to the pre-intervention transmission rate *β*_0_ multiplied by a scaling factor *b*_q_. Low values of *b*_q_ represent a substantial reduction in the transmission rate due to interventions. Social distancing guidelines were issued by New York City starting on March 17(62, 63), and a stay-at home order was issued which took effect on the evening of March 22(64). Thus,prior to the imposition of social distancing, the transmission rate of symptomatic individuals in our models, *β*(*t*), is equal to *β*_0_. From March 18th thru March 22nd, *β*(*t*) decreases linearly from *β*_0_ to *β*_1_. From March 23rd onwards, *β*(*t*) is equal to *β*_1_.

In all models, a fraction *p*_*S*_ of exposed individuals *E*_*m*_ becomes symptomatic. After an average of 5 days of transmission, symptomatic cases are hospitalized with probability *p*_H_. Symptomatic cases that are not severe enough to require hospitalization recover at rate *γ*. Hospitalized individuals recover at rate 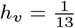 (30) and do not transmit while hospitalized. We assume a fixed population size for New York City of 8 million individuals (65).

Assumptions about which infected classes are infectious and how they contribute to the transmission rate allow us to reduce the SEPIAR model to the SEPIR or SEIAR models. Pre-symptomatic individuals transmit for an average of about a day (0.92 days(25)) at a transmission rate equal to the baseline transmission rate *β*(*t*) multiplied by a scaling factor b=*b*_p_. Asymptomatic infections transmit for an average of 5 days, equal to the average duration between the onset of symptoms and hospitalization in severe cases, at a transmission rate equal to the baseline rate *β*(*t*) multiplied by scaling factor *b*_a_.

The models are implemented numerically via an Euler approximation of the deterministic equations to which demographic stochasticity is added. Specifically, the number of individuals making state transitions from compartments with more than one exit is drawn from an Euler-multinomial distribution(66). The number of individuals making state transitions from compartments with only one exit is drawn from a binomial distribution.

### Description of Testing Model

The model takes into account daily changes in the testing capacity using estimates of daily tests conducted in New York City from the New York State Department of Health(23), as well as the re-testing of severe and non-severe symptomatic cases prior to leaving the hospital or quarantine. We assume that there are two categories of cases-severe (hospitalized) cases and non-severe cases subject to different testing priorities(67): the initial testing of new hospitalized COVID-19 cases (highest priority), the re-testing of those individuals when they leave the hospital, the testing of new non-severe symptomatic COVID-19 cases, and finally the re-testing of those symptomatic cases (lowest priority). All severe COVID-19 cases after March 1st are sampled when they enter the hospital and eventually tested once enough capacity is available. We assume that symptomatic non-severe cases are sampled at the same time in the course of their infection as severe cases. However, we assume that they are not tested if they recover before enough testing capacity is available. During the early stages of the epidemic, the CDC recommended test-based strategies to determine when to conclude home isolation or hospitalization(68). Accordingly, we assume that hospitalized cases are re-tested twice (over a 24 hour period) after the average length of time in the hospital (13 days), while non-severe cases are likewise re-tested twice after the end of a 14-day quarantine period.

We also take into account the potential for non-COVID-19 severe respiratory cases to be sampled in hospitals and tested (with the same priority as hospitalized COVID-19 cases). We use confirmed influenza cases(69) and syndrome surveillance reports of respiratory disease from emergency departments in New York City hospitals in previous years(70) to estimate the number of non-COVID-19 severe respiratory cases that may have been sampled (see SI Appendix). We assume that the RT-PCR testing has a sensitivity of 90%(71), that testing takes 48 hours(72), and that there is an additional negative-binomial distributed dispersion after the RT-PCR testing with standard deviation *σ*_M_. This dispersion takes into account variation in sampling and testing protocols across laboratories and hospitals, as well as variation in the sensitivity and time required for different PCR assays.

### Overview of the model fitting and inference strategy

Unless otherwise mentioned, we fit the following parameters: the recovery rate for non-severe symptomatic infections (*γ*), the scaling factors for asymptomatic, pre-symptomatic, and post-intervention transmission (*b*_a_, *b*_p_, and *b*_q_), the symptomatic probability (*p*_S_) and the hospitalization probability (*p*_H_), the reproductive number for symptomatic cases (*R*_0_), the dispersion parameter for RT-PCR testing (*σ*_*M*_), and the initial number of infected (*I*_0_) and exposed (*E*_0_) individuals at the start of the simulation on March 1, 2020. We use the iterated filtering algorithm MIF(73) within the R-package POMP (for partially observed Markov process models) to fit parameter combinations by likelihood maximization. We apply the Sequential Monte Carlo algorithm pfilter(74) to evaluate the likelihood of the final parameter combinations. In p articular, f or the analysis of the full SEPIAR model, we generate a Monte Carlo profile(75) for the relative strength of asymptomatic transmission (*b*_a_). For all resulting parameter combinations within 2 log–likelihood units of the MLE, we then calculate the likelihood with respect to serology using seroprevalence data from (24). We assume that each serology measurement is drawn from a binomial distribution with sample size N and proportion p equal to the observed seroprevalence. We isolate all combinations supported by the serology data that have log-likelihoods within 2 units of the MLE.

For each combination, we examine the proportion of cases that are symptomatic *p*_*S*_, the reproductive number in symptomatic individuals *R*_0_, and the overall reproductive number for the model 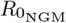. We derive the following expression for 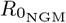 using the Next Generation Matrix(76) :

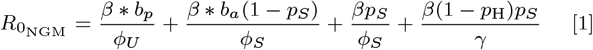

### Additional details

Further details of the SEPIAR equations, testing model, Monte Carlo Profile of the SEPIAR model, initial grid searches and model comparison of the SEPIR and SEIAR models, and derivation of the overall reproductive number 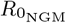, are provided in the SI Appendix.

## Data Availability

Original data created for the study are or will be available in a persistent repository upon publication.

## ACKNOWLEDGMENTS

R.S. was supported by a National Science Foundation Research Traineeship (no. 1735359: NRT-INFEWS: Computational data science to advance research at the energy environment nexus). The authors would like to thank Aaron King for his insightful discussions. This work was completed with resources and support provided by the University of Chicago’s Research Computing Center.

