## Supporting Information-Appendix for "Quantifying Asymptomatic Infection and Transmission of COVID-19 in New York City using Observed Cases, Serology and Testing Capacity"

September 18, 2020

**5 Overall Reproductive Number Derivation 23**

**6 Supplemental Figures 25**

### 1 SEPIAR Model Details

#### 1.1 SEPIAR Model Compartments

#### 1.2 ODE Equations

$$\frac{dS}{dt} = -(\lambda_{\mathrm{FOI}}(t))S(t) \tag{1}$$

For exposed compartment $E_m$ where $m = 1$:

$$\frac{dE_1}{dt} = (\lambda_{\mathrm{FOI}}(t))S(t) - \phi_{\mathrm{E}} E_1(t) \tag{2}$$

| **Compartment** | **Infection Status** |
|---|---|
| $S(t)$ | Susceptible |
| $E_m(t)$ | Exposed in compartment $m$ |
| $P(t)$ | Pre-symptomatic |
| $A(t)$ | Asymptomatic |
| $I_{\mathrm{S}_1}(t)$ | Infected symptomatic not yet sampled |
| $I_{\mathrm{S}_2}(t)$ | Infected symptomatic (non-severe) |
| $H(t)$ | Hospitalized (severe infected) |
| $R_{\mathrm{A}}(t)$ | Recovered (Asymptomatic) |
| $R_{\mathrm{F}}(t)$ | Recovered (Symptomatic Non-Severe) |
| $R_{\mathrm{H}}(t)$ | Recovered (Severe) |

Table 1: Comparmtents in the SEPIAR epidemiological model

For exposed compartment $E_m$ where $1 < m <= M$:

$$\frac{dE_m}{dt} = \phi_{\mathrm{E}} E_{m-1}(t) - \phi_{\mathrm{E}} E_m(t) \tag{3}$$

$$\frac{dP}{dt} = \phi_{\mathrm{E}} E_M(t) - \phi_{\mathrm{U}} P(t) \tag{4}$$

$$\frac{dI_{\mathrm{S}_1}}{dt} = p_{\mathrm{S}} \phi_{\mathrm{U}} P(t) - \phi_{\mathrm{S}} I_{\mathrm{S}_1}(t) \tag{5}$$

$$\frac{dH}{dt} = p_{\mathrm{H}} \phi_{\mathrm{S}} I_{\mathrm{S}_1}(t) - h_{\mathrm{V}} H(t) \tag{6}$$

$$\frac{dI_{\mathrm{S}_2}}{dt} = (1 - p_{\mathrm{H}}) \phi_{\mathrm{S}} I_{\mathrm{S}_1}(t) - \gamma I_{\mathrm{S}_2}(t) \tag{7}$$

$$\frac{dA}{dt} = (1 - p_{\mathrm{S}}) \phi_{\mathrm{U}} P(t) - \phi_{\mathrm{S}} A \tag{8}$$

$$\frac{dR_{\mathrm{A}}}{dt} = \phi_{\mathrm{S}} A \tag{9}$$

$$\frac{dR_{\mathrm{F}}}{dt} = \gamma I_{\mathrm{S}_2}(t) \tag{10}$$

$$\frac{dR_{\mathrm{H}}}{dt} = h_{\mathrm{V}} H(t) \tag{11}$$

$$\lambda_{\mathrm{FOI}}(t) = \frac{\beta(t)[(I_{\mathrm{S}_1}(t)) + (I_{\mathrm{S}_2}(t))] + \beta_{\mathrm{a}}(t)[A(t)] + \beta_{\mathrm{p}}(t)[P(t)]}{N} \tag{12}$$

##### 1.3 Accumulator Variables

Let $C_{\mathrm{Q1}}$ represent the total number of individuals with severe COVID-19 cases who enter the hospital over a single-day period. In the SEPIAR model, this is the number of people moving from compartment $I_{\mathrm{S}_1}$ to $H$ in a single day.

We assume that non-severe COVID-19 cases are sampled at the same time in the course of their infection as severe cases, provided that sufficient testing capacity is available. Let $C_{\mathrm{Q3}}$ represent the total number of people who move from compartment $I_{\mathrm{S}_1}$ to compartment $I_{\mathrm{S}_2}$ over a single day in the SEPIAR model. These people represents symptomatic COVID-19 cases that do not become severe.

The quantities $C_{\mathrm{Q1}}$ and $C_{\mathrm{Q3}}$ generated from the epidemiological model are used as inputs for the testing model.

#### 2 Model Fitting Techniques

##### 2.1 Fitted Parameters

Unless otherwise mentioned, we fit the recovery rate for non-severe symptomatic infections ($\gamma$), the scaling factors for asymptomatic and pre-symptomatic transmission ($b_{\mathrm{a}}$ and $b_{\mathrm{p}}$), the scaling factor for post-intervention transmission ($b_{\mathrm{q}}$), the proportion of new infected cases that will become symptomatic ($p_{\mathrm{S}}$), the proportion of symptomatic cases that are severe enough to require hospitalization ($p_{\mathrm{H}}$), the reproductive number for symptomatic cases ($R_0$) the dispersion parameter for RT-PCR testing ($\sigma_{\mathrm{M}}$), and the initial number of infected ($I_0$) and exposed ($E_0$) individuals at the start of the simulation on March 1, 2020. We constrain the fitting algorithm to explore only positive values for all fitted parameters and only values between 0 and 1 for $p_{\mathrm{S}}$, $p_{\mathrm{H}}$, $b_{\mathrm{a}}$, $b_{\mathrm{p}}$, and $b_{\mathrm{q}}$.

##### 2.2 Initial Grid Searches of SEPIR and SEIAR Models

We first fit the SEPIR model, which does not have asymptomatic transmission, and the SEIAR model, which does not have pre-sympamatic tranmission (Figure 11). For each model, we generate a grid of 25,000 initial parameter combinations using Latin Hypercule Sampling. For each initial combination, the given model is fit to observed case data for two sets of 50 iterations using the iterated filtering algorithm MIF2[1] using 50,000 particles. The final likelihood of each parameter combination with respect to observed case data is then estimated using the sequential Monte Carlo algorithm pfilter[2]. We then isolate all parameter combinations within 2 log-likelihood units of the parameter combination with the highest likelihood (the maximum likelihood estimate or MLE), and calculate the likelihood of each combination with respect to the serology data. We then isolate the parameter combination with the highest likelihood with respect to the serology data.

##### 2.3 Monte Carlo Profile of SEPIAR Model

For the analysis of the full SEPIAR model, which includes both pre-symptomatic and asymptomatic transmission, we construct a Monte Carlo Profile[3] of the relative strength of asymptomatic transmission ($b_{\mathrm{a}}$). As Figure 2 illustrates, we generate a set of starting points at 30 different evenly spaced values for $b_{\mathrm{a}}$

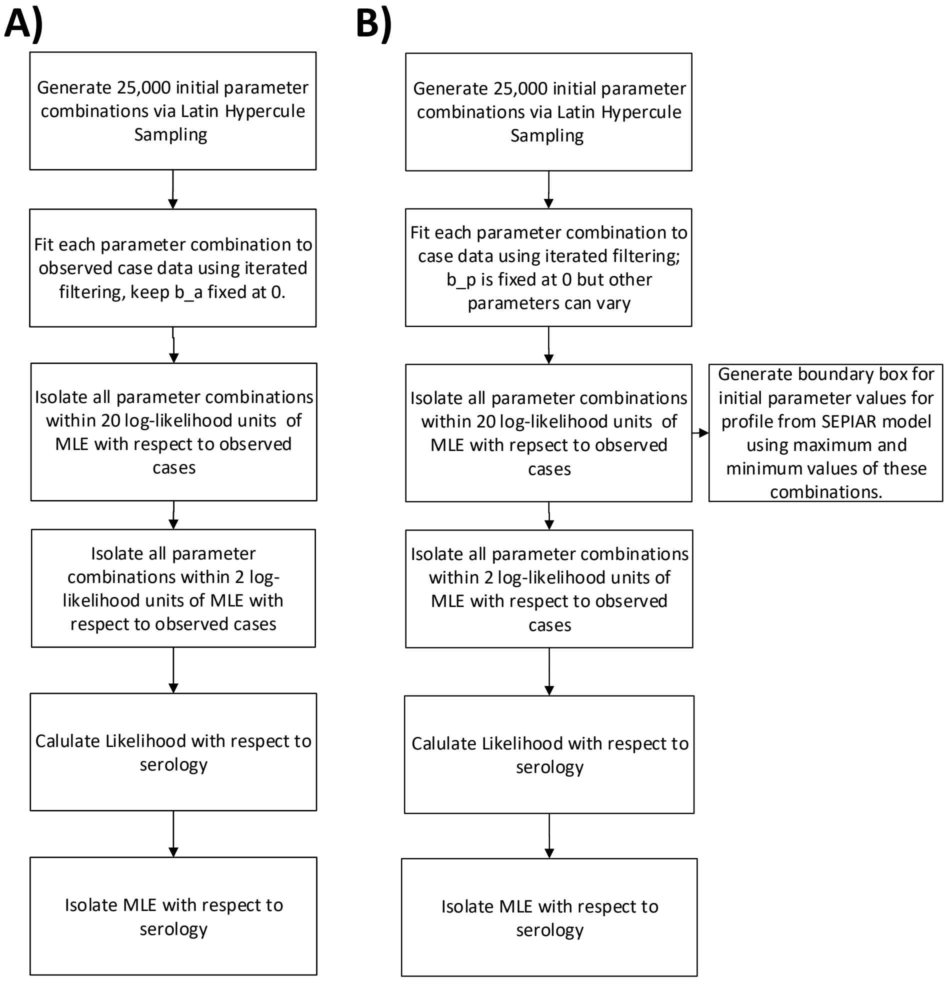

Figure 1: **Diagram of the grid searches for the SEPIR (A) and SEIAR (B) models.**

between 0 and 1. For each of those 30 starting points, we create 40 different initial sampling points with the same value of $b_{\mathrm{a}}$ but different values for the other parameters being fitted. Initial values for the relative strength of pre-symptomatic transmission $b_p$ were drawn from a uniform distribution between 0 and 1. The values for all of the other fitted parameters were uniformly drawn from the boundaries of all parameter combinations obtained from fitting the SEIAR model that had likelihoods with respect to case data within 20 log-likelihood units of the MLE. This yielded a total of 1200 starting points. Each profile starting point was then fit to case data using the iterated filtering algorithm MIF2[1] and the Sequential Monte Carlo algorithm pfilter[2], with MIF2 constrained to keep $b_{\mathrm{a}}$ unchanged. For all parameter combinations that were supported by observed case data (i.e. that had log-likelihoods within 2 units of the MLE), we then calculated the likelihood with respect to serology. All parameter combinations from the full SEPIAR model with serology likelihoods within 2-log-likelihood units of the MLE were used in subsequent analyses of the proportion of cases that are symptomatic ($p_{\mathrm{S}}$), the reproductive number in symptomatic individuals ( $R_0$), and the overall reproductive number for the model ($R_{0_{\mathrm{NGM}}}$) which was calculated using the Next Generation Matrix[4].

#### 2.4 Model Comparison

We compare the likelihoods with respect to the serology data of all the maximum likelihood estimates from the SEPIR, SEIAR, and SEPIAR models via the Likelihood Ratio Test. Recall that when calculating the likelihood with respect to case data, all three models had maximum likelihoods that were not statistically different.

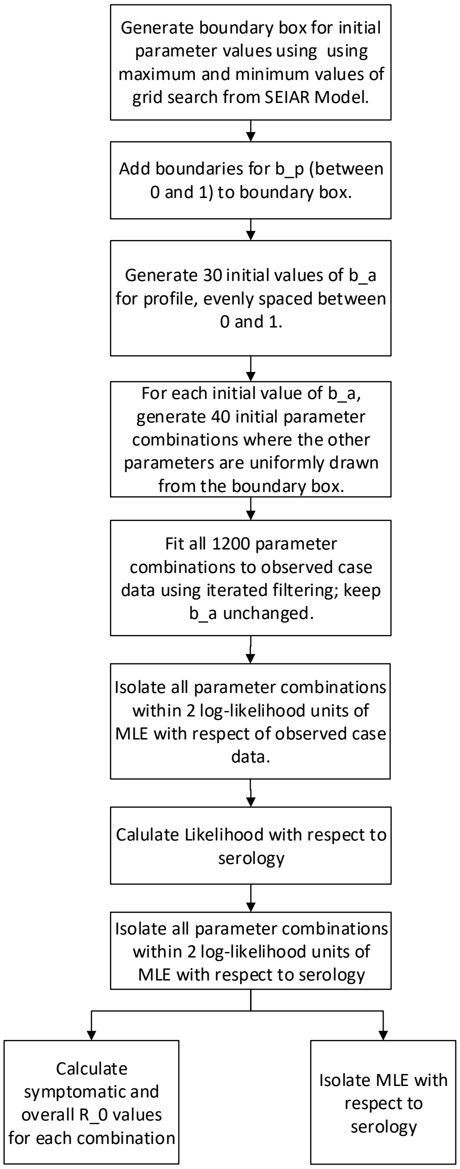

Figure 2: **SEPIAR Profile Fitting Procedure**. This diagram summarizes how the Monte Carlo profile of $b_a$ for the SEPIAR model was fit to case data and subsequently to serology.

| Model | Log-Likelihood |
|---|---|
| SEPIAR | -24.34239** |
| SEIAR | -24.6274** |
| SEPIR | -29.17705 |

Table 2: Comparison of MLE Likelihoods from SEPIR, SEIAR, and SEPIAR models with respect to serology data

### 3 Testing Model

The testing model is implemented with discrete time steps of a day, denoted hereafter by $\mathbf{t}$.

#### 3.1 Testing Capacity Data

We use the total number of RT-PCR tests conducted each day across the five counties compromising New York City as measured by the New York State Health Department's COVID tracker as the daily testing capacity for the city. This capacity is fed into the model as a co-variate ($L_{\text{reported}}(\mathbf{t})$).

Let $L_0(\mathbf{t})$ represent the initial testing capacity on day $\mathbf{t}$. Recall that we assume it took 2 days to conduct a RT-PCR Test. Therefore, we advance the testing capacity by 2 days from the observed value, since the testing capacity on day $\mathbf{t}$ will correspond to the number of tests conducted on day $\mathbf{t}+2$.

$$L_0(\mathbf{t}) = L_{\text{reported}}(\mathbf{t}+2) \tag{13}$$

#### 3.2 Testing Priorities

Given a finite daily testing capacity, we assume that tests are used in the following order during any given day until the testing capacity is exhausted:

1. **Initial testing of hospitalized patients.** Hospitalized patients include patients who have severe cases of COVID-19 (Queue 1) as well as indi-

viduals who do not have COVID-19 but are hospitalized with respiratory symptoms (Queue NC). Patients are added to queue 1 as they enter compartment $H$ in the epidemiological model.

2. **Re-testing hospitalized patients when they leave the hospital (Queue 2).** Patients hospitalized with COVID-19 are re-tested twice over a 24 hour period as they exit the hospital.

3. **Initial testing of non-severe symptomatic cases (Queue 3)**.Patients are added to queue 1 as they enter compartment $I_{S_2}$ in the epidemiological model.

4. **Re-testing of non-severe symptomatic cases 14 days after they are first sampled (Queue 4).**

We assume that any remaining unused testing capacity remaining after all four queues have been tested is used to test individuals without COVID-19. Thus, this unused capacity does not roll over to the next day.

Supporting Figure S4 illustrates these testing priorities.

#### 3.3 General framework for incorporating changes in testing capacity

Below we provide the detailed steps of the general testing framework which can be applied to other locations where testing capacity is changing over time. A diagram of this framework is shown in Supporting Figure S3. We illustrate the steps by focusing on the testing of hospitalized severe COVID-19 cases. In subsection 3.4, we describe several modifications to this framework that we make to take into account additional queues and modifications that are specific to New York City in the early stage of the epidemic.

We use the variable $Q_1$ to represent all COVID-19 hospitalized cases that have been sampled but have not been tested yet. We first add the hospitalized COVID-19 cases that have just been sampled on day $\mathbf{t}$ ($C_{Q1}(\mathbf{t})$) to $Q_1$.

$$Q_{1_1}(\mathbf{t}) = Q_{1_1}(\mathbf{t}) + C_{Q1}(\mathbf{t}) \tag{14}$$

Let the variable $T_{\mathrm{Q1}}$ represent all of the people in Queue 1 who will be tested. If $L_0(\mathbf{t})$ is bigger than the Queue 1, then everyone in Queue 1 can be tested and

$$T_{\mathrm{Q1}}(\mathbf{t}) = Q_1(\mathbf{t}) \tag{15}$$

Let $L_{\mathrm{unused}}(\mathbf{t})$ represent the testing capacity left over after Q1 has been tested:

$$L_{\mathrm{unused}}(\mathbf{t}) = L_0(\mathbf{t}) - T_{\mathrm{Q1}}(\mathbf{t}) \tag{16}$$

There are then no individuals left in Q1 who need to be tested.

$$Q_1(\mathbf{t}) = 0 \tag{17}$$

However, suppose that on a given day there is not enough testing capacity to test everyone in Q1 (i.e. at the start of that day, $L_0(\mathbf{t}) < Q_1(\mathbf{t})$).

In this case, we assume that all of the testing capacity is used to test Q1:

$$T_{\mathrm{Q1}}(\mathbf{t}) = L_0(\mathbf{t}) \tag{18}$$

We therefore decrease Queue 1 by the number of people who were tested.

$$Q_1(\mathbf{t}) = Q_1(\mathbf{t}) - T_{\mathrm{Q1}}(\mathbf{t}) \tag{19}$$

In this case, there is no unused testing capacity:

$$L_{\text{unused}}(\mathbf{t}) = 0 \tag{20}$$

Let $Y_{\text{Q1}}(\mathbf{t})$ represent the number of people who tested positive on day $\mathbf{t}$, and $c$ the sensitivity of the RT-PCR assay.

We simulate RT-PCR testing as a draw from a Binomial distribution where $N = T_{\text{Q1}}(\mathbf{t})$ and $p = c$.

$$Y_{\text{Q1}}(\mathbf{t}) \sim Binomial(T_{\text{Q1}}(\mathbf{t}), c) \tag{21}$$

If there is unused testing capacity after everyone in Queue 1 has been tested (i.e. $L_{\text{unused}}(\mathbf{t}) > 0$, then this capacity can be used to test individuals in other queues with a lower priority; see Section 3.4).

If testing of any of the individuals in these later queues results in additional new positive cases, these are added to $Y_{\text{Q1}}(\mathbf{t})$ to obtain the total number of expected new COVID-19 cases during day $\mathbf{t}$, denoted by $Y_{\text{sum}}(\mathbf{t})$.

#### 3.4 NYC Specific Modifications and Additional Queues

There are several aspects of the testing model that are particular to New York City and the initial wave of the epidemic in the U.S. We summarize those aspects here. With sufficient description of the specific testing implementation, similar modifications could be considered for other locations.

- **Re-Sampling of Hospitalized Cases-** We assume that hospitalized COVID-19 cases will be re-sampled twice over a 24-hour period as they leave the hospital, to make sure that they have recovered. We take this into account with a second queue, Queue 2 ($Q_2(\mathbf{t})$), which represents patients who tested positive who had a severe COVID-19 infection, were re-sampled

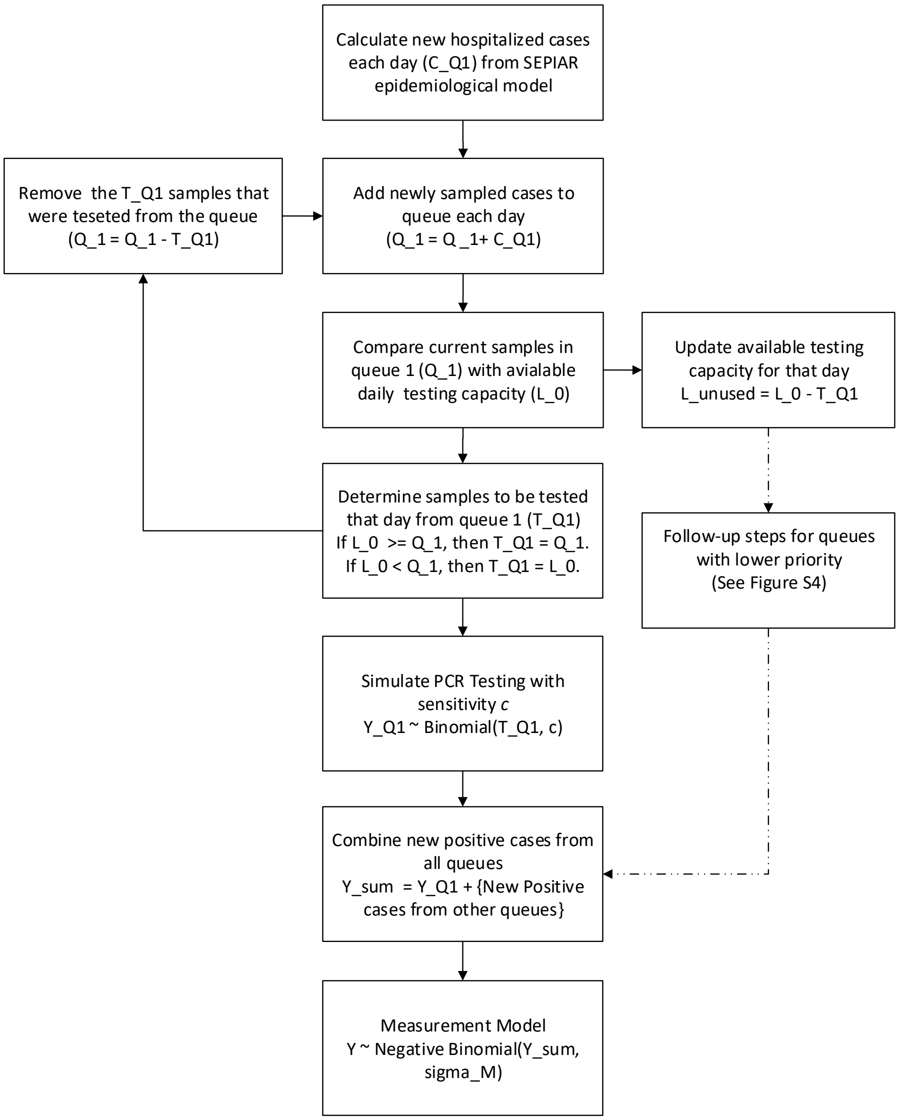

Figure 3: **Diagram of the general testing framework described in Section 3.3**. The New York City-specific modifications described in Section 3.4 are not shown here.

as they left the hospital, and need to be tested again. We do not simulate RT-PCR testing, keep track of results of testing, or deal with lags for Queue 2, since the RT-PCR results of this re-sampling are not relevant for our model. What is relevant is that at least some of the available testing capacity $L_{\text{unused}}(\mathbf{t})$ may be used up re-testing hospitalized patients as they leave the hospital before other groups such as non-severe symptomatic patients can be tested.

In our epidemiological model, we assume that individuals spend an average of 13 days in the hospital. To be consistent, in our testing model, we assume that individuals who test positive for COVID-19 will be re-sampled once 13 days after they enter Queue 1, and then re-sampled a second time 1 day later. To keep track of the days since each sample entered Queue 1, we modify our implementation of Queue 1 by introducing initial sampling cohorts. There are thirteen cohorts, representing people who entered Queue 1 up to 13 days before time $\mathbf{t}$. Let $T_{\text{Q1}_v}(\mathbf{t})$ represent the number of people who were sampled $v$ days before day $\mathbf{t}$ who will be tested on day $\mathbf{t}$. We calculate $T_{\text{Q1}_v}(\mathbf{t})$ as we loop through each cohort $v$ in Queue 1, $Q_{1_v}$, starting with the oldest ($Q_{1_V}$) and ending with the most recent ($Q_{1_1}$).

For example, if there is sufficient capacity to test everyone in the oldest cohort (i.e. $L_0 > Q_{1_V}(\mathbf{t})$), then we essentially follow a similar procedure to equations 15 and 16:

$$T_{\text{Q1}_V}(\mathbf{t}) = Q_{1_V}(\mathbf{t}) \tag{22}$$

$$L_{\text{unused}}(\mathbf{t}) = L_0(\mathbf{t}) - T_{\text{Q1}_V}(\mathbf{t}) \tag{23}$$

We similarly decrease $L_{\text{unused}}(\mathbf{t})$ as the capacity is used up from testing each subsequent cohort. For example, suppose that there is enough capacity to test a subsequent cohort $v$ (i.e. $L_{\text{unused}}(\mathbf{t}) > Q_{1_v}(\mathbf{t})$). Then:

$$T_{\text{Q1}_v}(\mathbf{t}) = Q_{1_v}(\mathbf{t}) \tag{24}$$

$$L_{\text{unused}}(\mathbf{t}) = L_{\text{unused}}(\mathbf{t}) - T_{\text{Q1}_v}(\mathbf{t}) \tag{25}$$

Alternatively, when we do not have enough testing capacity to test all of the cohort, then:

$$T_{\text{Q1}_v}(\mathbf{t}) = L_{\text{unused}}(\mathbf{t}) \tag{26}$$

We loop through all cohorts in Queue 1 until either all the people in the queue have been tested or until the unused testing capacity is exhausted. When simulating RT-PCR testing, we again loop over all sampling cohorts $v$ from $1 : V$. Equation 21 is modified accordingly:

$$Y_{\text{Q1}_v}(\mathbf{t}) \sim Binomial(T_{\text{Q1}_v}(\mathbf{t}), c) \tag{27}$$

For the first re-sampling, we need to keep track of those individuals who tested positive when they first entered the hospital. Because different cases form the same cohort may be tested on different days, we need a variable to accumulate those cases that tested positive and belong to the same cohort. Let $P_{\text{Q1}_v}(\mathbf{t})$ represent all cases from Queue 1 initially sampled $v - 1$ days before time $\mathbf{t}$ who have tested positive so far.

We accumulate the total number of people in initial sampling cohort $v$ who have tested positive so far by adding the number of people from that cohort who tested positive on day $\mathbf{t}$ ($Y_{\mathrm{Q1}_v}(\mathbf{t})$) to $P_{\mathrm{Q1}_v}(\mathbf{t})$:

$$P_{\mathrm{Q1}_v}(\mathbf{t}) = P_{\mathrm{Q1}_v}(\mathbf{t}) + Y_{\mathrm{Q1}_v}(\mathbf{t}) \tag{28}$$

For the first re-sampling, the oldest cohort is entered into $Q_2(\mathbf{t})$ :

$$Q_2(\mathbf{t}) = Q_2(\mathbf{t}) + P_{\mathrm{Q1}_V}(\mathbf{t}) \tag{29}$$

For the second re-sampling, we keep track of the individuals in this oldest cohort and enter them into Q2 again one day later (on day $t + 1$).

At the end of each simulation day, all initial sampling cohorts are advanced by 1 day.

- **2-Day Lag in Testing-** We incorporate a 2-day lag between when tests are conducted and results are reported to take into account the 48 hour testing time of early RT-PCR tests. We do not update cohorts during this lag, so this effectively adds another 2 days between the initial sampling and the re-sampling beyond the 13 days spent in the hospital. If this framework is applied to other locations and time periods, this modification may not be needed, particularly if rapid diagnosis RT-PCR tests are in use.

- **Testing of non-symptomatic severe cases-**Queue 3, which records the number of non-severe symptomatic cases that need to be tested, is implemented identically to Queue 1, except that individuals can exit the queue at rate $\gamma$ as they recover, even if they have not yet been tested. Sampling cohorts are used in this queue as well.

- **Re-testing of non-symptomatic severe cases-** Early CDC guidelines[5] recommended a 14-day quarantine for non-hospitalized symptomatic individuals, and that these individuals be re-tested at the end of the quarantine. Queue 4, which records the re-sampling of non-severe symptomatic cases as they exit quarantine, is implemented identically to Queue 2, except that cases are re-sampled 14 days after they enter Queue 3, rather than 13 days, to match the length of the quarantine period.

- **Queues are numbered in order of priority-** Any unused testing capacity after Queue 1 is empty is applied to Queue 2, and subsequently Queues 3 and 4.

- **Severe non-COVID hospitalized cases-** Queue NC, which represents severe non-COVID-19 respiratory cases, is implemented in a similar fashion as Queue 1. Let $G_{\text{severe}}(w, y)$ represent our estimate of the weekly expected severe non-COVID-19 respiratory cases that would be sampled for testing in the hospital, where $w$ denotes the week, and $y$, the year. This estimate is obtained from syndrome surveillance data as described later in Section 4. The number of daily new cases that enter Queue NC, $C_{\text{Q}_{\text{NC}}}$, is therefore:

$$C_{\text{Q}_{\text{NC}}} = \frac{G_{\text{severe}}(w, y)}{7} \tag{30}$$

Queue NC has the same priority as Queue 1, since both groups of individuals will present with severe respiratory symptoms at the time of sampling. Both groups are hence sampled at once. For the situation where testing capacity is greater than the total samples in a given sampling cohort in Queue 1 and Queue NC, we simply test all samples from that cohort in both queues. For the situation where the testing capacity is limiting, we simulate a draw from a hypergeometric distribution. For example, in the model, we modify equation 26 as follows:

$$T_{\text{Q1}_v}(\mathbf{t}) \sim \textit{hypergeom}(Q_{1_v}(\mathbf{t}), Q_{\text{NC}_v}(\mathbf{t}), L_{\text{unused}}(\mathbf{t})) \tag{31}$$

We assume that the RT-PCR test is 100% specific, so all cases in Queue NC will test negative since they do not have COVID-19. We thus do not simulate RT-PCR testing for Queue NC. The main importance of individuals in Queue NC is that they deplete some of the testing capacity that would otherwise be used for Queue 1.

##### 3.5 Measurement Model

Let $Y_{\text{sum}}$ represent the total number of new positive tests from all of the queues.

We assume that there is additional negative-binomial distributed dispersion after the RT-PCR testing with standard deviation $\sigma_{\text{M}}$. Thus, if $Y$ is the observed number number of daily cases, we simulate $Y$ from a negative binomial distribution with mean equal to $Y_{\text{sum}}$ and and variance $Y_{\text{sum}} + \frac{\sigma_{\text{M}}^2}{{Y_{\text{sum}}}^2}$

#### 4 Syndrome Surveillance Estimates

We estimate the number of non-COVID-19 severe respiratory cases that may have presented each week of the epidemic in NYC hospitals using syndrome surveillance data from NYC hospital emergency departments and observed influenza cases in NYC in previous years. The estimate we seek here represents the typical number of non-COVID-19 respiratory cases one would expect in a given week of the year given the seasonal pattern of influenza cases and that of other respiratory ailments that present to NYC hospitals.

##### 4.1 Description of Data

###### 4.1.1 Syndrome Surveillance for Respiratory Disease

Weekly respiratory syndrome surveillance reports for all emergency departments in New York City hospitals from 2016-2020 were obtained from the New York

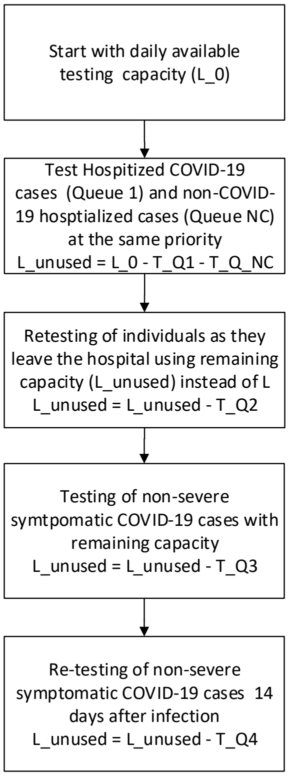

Figure 4: **Diagram of the testing priorities described in Section 3.2.**

Health Department's web portal[6]. These reports include all cases in which the chief complaint mentioned bronchitis, chest cold, chest congestion, chest pain, cough, difficulty breathing, pneumonia, shortness of breath, or upper respiratory infection. For this analysis, we use weeks 2-20 from 2016,2017 and 2019.

###### 4.1.2 Confirmed Flu Cases

Confirmed influenza cases from all New York Counties from 2016-2020 for all counties in New York State were obtained from the New York State public health portal[7]. We used data from the five countries comprising New York City that correspond to the same time period as the syndrome surveillance data. We excluded 2018 from this analysis since 2018 was an anomalous, severe, influenza season[8].

##### 4.2 Description of Statistical Model

Recall that $G_{\text{severe}}(w, y)$ is the number of non-COVID-19 respiratory infections during week $w$ of year $y$ that were severe enough to be sampled for COVID-19 testing. This was the quantity added to Queue NC in Equation 30 of Section 3.4. We assume that these cases are a fixed fraction $s$ of the total non-COVID respiratory syndrome cases presenting in the emergency departments of hospitals in NYC in week $w$ of year $y$, which we denote as $(G(w, y))$:

$$G_{\text{severe}}(w, y) = G(w, y) * s \tag{32}$$

We assume that in the absence of COVID-19, a portion of observed respiratory syndrome surveillance reports are associated with influenza, and that the non-influenza associated reports have a fixed seasonality.

Therefore, we consider that our estimate for the non-COVID-19 weekly respiratory syndrome surveillance cases $(G(w, y))$ varies linearly with confirmed influenza cases $F(w, y)$ in NYC and presents additional weekly variability whose effect is represented non-para-metrically with a polynomial dependency as follows:

$$G(w, y) = g_0 + g_F F(w, y) + b_3 w^3 + b_2 w^2 + b_1 w + b_0 + \epsilon \tag{33}$$

where

$$\epsilon \sim rnorm(0, \sigma_\epsilon) \tag{34}$$

###### 4.2.1 Model Fitting and Simulation

We estimate the intercept $g_0$ and influenza coefficient $g_F$ via a linear regression of respiratory syndrome surveillance reports and confirmed cases in New York City in 2016,2017 and 2019. We then fit the residuals from this linear regression to a third-order polynomial seasonality function to obtain estimates of coefficients $b_i$. We estimate the error term $\sigma_\epsilon$ by measuring the residual standard error from the polynomial fit.

When fitting the epidemiological model, we simulate values for $G(w, y)$ using observed weekly influenza cases in 2020 as the co-variate $F(w, y)$ obtained from the same source[7]. A plot of the fitted model is shown in Figure S11.

##### 4.3 Estimation of Proportion Sampled for COVID-19 testing

Recall that the scaling parameter $s$ represents the probability that an individual who shows up to the emergency department with respiratory symptoms is severe enough to merit testing for COVID-19. As a proxy for this value, we use the ratio of individuals aged 65 or older who were hospitalized for influenza to the number of individuals aged 65 or older who had a medical visit for influenza during the 2018-2019 influenza season[9].

The value we use for this scaling parameter $s$ for the fitting of all three models is 0.16.

We profile over this parameter value as a sensitivity analysis(see Figure S9). Allowing this parameter to vary does not result in a higher likelihood with respect to serology data compared to the MLE parameter combination from the main analysis.

##### 4.3.1 Table of Fitted Parameters

| **Parameter** | **Value** |
|---|---|
| $g_{\mathrm{F}}$ | 0.12 |
| $g_0$ | 1183 |
| $b_3$ | 0.012 |
| $b_2$ | 0.981 |
| $b_1$ | -37.2 |
| $b_0$ | 229 |
| $\sigma_\epsilon$ | 109 |
| $s$ | 0.16 |

Table 3: Parameter values used for the non-COVID-19 severe cases estimate (rounded).

#### 4.4 Detection of anomalies in 2020 syndrome surveillance

From our model of non COVID-19 respiratory cases, we can obtain an estimate of the expected number of syndrome surveillance reports $G_{w,y}$ in 2020 in the absence of COVID-19. Note that we do not use the scaling parameter $s$ in this calculation, unlike when fitting the epidemiological model.

If we subtract this value from the observed respiratory syndrome surveillance reports in 2020, we obtain a metric for anomalous respiratory syndrome surveillance reports related to COVID-19. This is the pink line in Figure S5.

#### 5 Overall Reproductive Number Derivation

Following [4], we derive $R_{0_{\mathrm{NGM}}}$ as the leading eigenvalue of the following matrix:

$$K = -T\Sigma^{-1}, \tag{35}$$

which is composed of two other matrices, $T$ and $\Sigma^{-1}$, defined below.

The Transmission Matrix is given by

$$T = \begin{matrix} 0 & 0 & 0 & 0 & 0 & \beta_P & \beta_A & \beta & \beta \\ 0 & 0 & 0 & 0 & 0 & 0 & 0 & 0 & 0 \\ 0 & 0 & 0 & 0 & 0 & 0 & 0 & 0 & 0 \\ 0 & 0 & 0 & 0 & 0 & 0 & 0 & 0 & 0 \\ 0 & 0 & 0 & 0 & 0 & 0 & 0 & 0 & 0 \\ 0 & 0 & 0 & 0 & 0 & 0 & 0 & 0 & 0 \\ 0 & 0 & 0 & 0 & 0 & 0 & 0 & 0 & 0 \\ 0 & 0 & 0 & 0 & 0 & 0 & 0 & 0 & 0 \\ 0 & 0 & 0 & 0 & 0 & 0 & 0 & 0 & 0 \end{matrix}$$

The Transition Matrix is given by

$$\Sigma = \begin{pmatrix} -\phi_E & 0 & 0 & 0 & 0 & 0 & 0 & 0 & 0 \\ \phi_E & -\phi_E & 0 & 0 & 0 & 0 & 0 & 0 & 0 \\ 0 & \phi_E & -\phi_E & 0 & 0 & 0 & 0 & 0 & 0 \\ 0 & 0 & \phi_E & -\phi_E & 0 & 0 & 0 & 0 & 0 \\ 0 & 0 & 0 & \phi_E & -\phi_E & 0 & 0 & 0 & 0 \\ 0 & 0 & 0 & 0 & \phi_E & -\phi_U & 0 & 0 & 0 \\ 0 & 0 & 0 & 0 & 0 & (1-p_S)\phi_U & -\phi_S & 0 & 0 \\ 0 & 0 & 0 & 0 & 0 & p_S\phi_U & 0 & -\phi_S & 0 \\ 0 & 0 & 0 & 0 & 0 & 0 & 0 & (1-p_{\mathrm{H}})\phi_S & -\gamma \end{pmatrix}$$

Then, the inverse of the transition matrix is:

$$\Sigma^{-1} = \begin{pmatrix} \frac{-1}{\phi_E} & 0 & 0 & 0 & 0 & 0 & 0 & 0 & 0 \\ \frac{-1}{\phi_E} & \frac{-1}{\phi_E} & 0 & 0 & 0 & 0 & 0 & 0 & 0 \\ \frac{-1}{\phi_E} & \frac{-1}{\phi_E} & \frac{-1}{\phi_E} & 0 & 0 & 0 & 0 & 0 & 0 \\ \frac{-1}{\phi_E} & \frac{-1}{\phi_E} & \frac{-1}{\phi_E} & \frac{-1}{\phi_E} & 0 & 0 & 0 & 0 & 0 \\ \frac{-1}{\phi_E} & \frac{-1}{\phi_E} & \frac{-1}{\phi_E} & \frac{-1}{\phi_E} & \frac{-1}{\phi_E} & 0 & 0 & 0 & 0 \\ \frac{-1}{\phi_U} & \frac{-1}{\phi_U} & \frac{-1}{\phi_U} & \frac{-1}{\phi_U} & \frac{-1}{\phi_U} & \frac{-1}{\phi_U} & 0 & 0 & 0 \\ \frac{-(1-p_S)}{\phi_S} & \frac{-(1-p_S)}{\phi_S} & \frac{-(1-p_S)}{\phi_S} & \frac{-(1-p_S)}{\phi_S} & \frac{-(1-p_S)}{\phi_S} & \frac{-(1-p_S)}{\phi_S} & \frac{-1}{\phi_S} & 0 & 0 \\ \frac{-p_S}{\phi_S} & \frac{-p_S}{\phi_S} & \frac{-p_S}{\phi_S} & \frac{-p_S}{\phi_S} & \frac{-p_S}{\phi_S} & \frac{-p_S}{\phi_S} & 0 & \frac{-1}{\phi_S} & 0 \\ \frac{-(1-p_{\mathrm{H}})p_S}{\gamma} & \frac{-(1-p_{\mathrm{H}})p_S}{\gamma} & \frac{-(1-p_{\mathrm{H}})p_S}{\gamma} & \frac{-(1-p_{\mathrm{H}})p_S}{\gamma} & \frac{-(1-p_{\mathrm{H}})p_S}{\gamma} & \frac{-(1-p_{\mathrm{H}})p_S}{\gamma} & 0 & \frac{-(1-p_{\mathrm{H}})}{\gamma} & \frac{-1}{\gamma} \end{pmatrix}$$

From the leading eignvalue of the resulting matrix K, we finally obtain:

$$R_{0_{NGM}} = \frac{\beta_P}{\phi_U} + \frac{\beta_A(1-p_S)}{\phi_S} + \frac{\beta p_S}{\phi_S} + \frac{\beta(1-p_{\mathrm{H}})p_S}{\gamma} \tag{36}$$

#### 6 Supplemental Figures

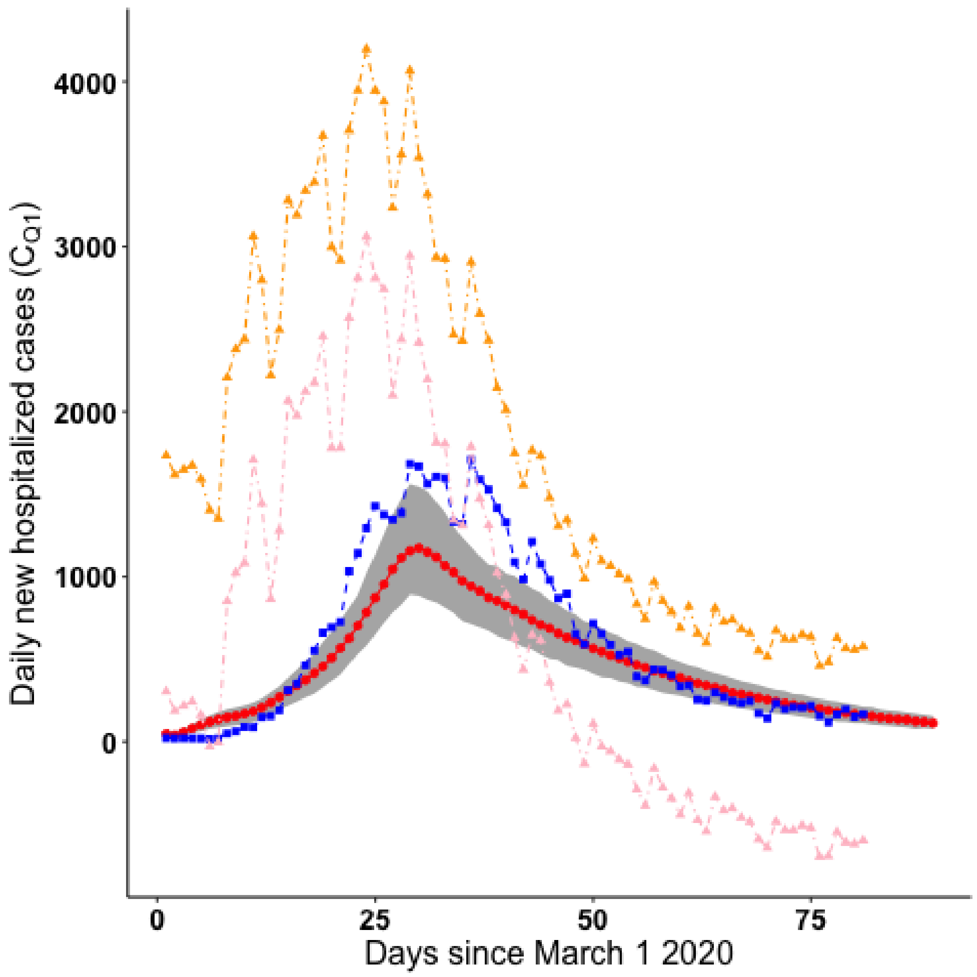

Figure 5: **Comparison of daily COVID hospitalizations under the model with observed COVID hospitalizations in New York City and emergency department respiratory syndrome surveillance reports.** The red line represents the median daily case hospitalizations from 100 simulations from the parameter combination with the highest likelihood with respect to serology from the $b_a$ profile. The grey shading represents the bounds of the 2.5% and 97.5% quantiles across all parameter combinations from the $b_a$ profile that are supported by case and serology data. The blue line shows observed COVID daily hospitalizations in New York City. The yellow line denotes daily reports of respiratory illness from syndrome surveillance in New York City emergency departments, while the pink line denotes anomalous respiratory surveillance reports compared to previous years.

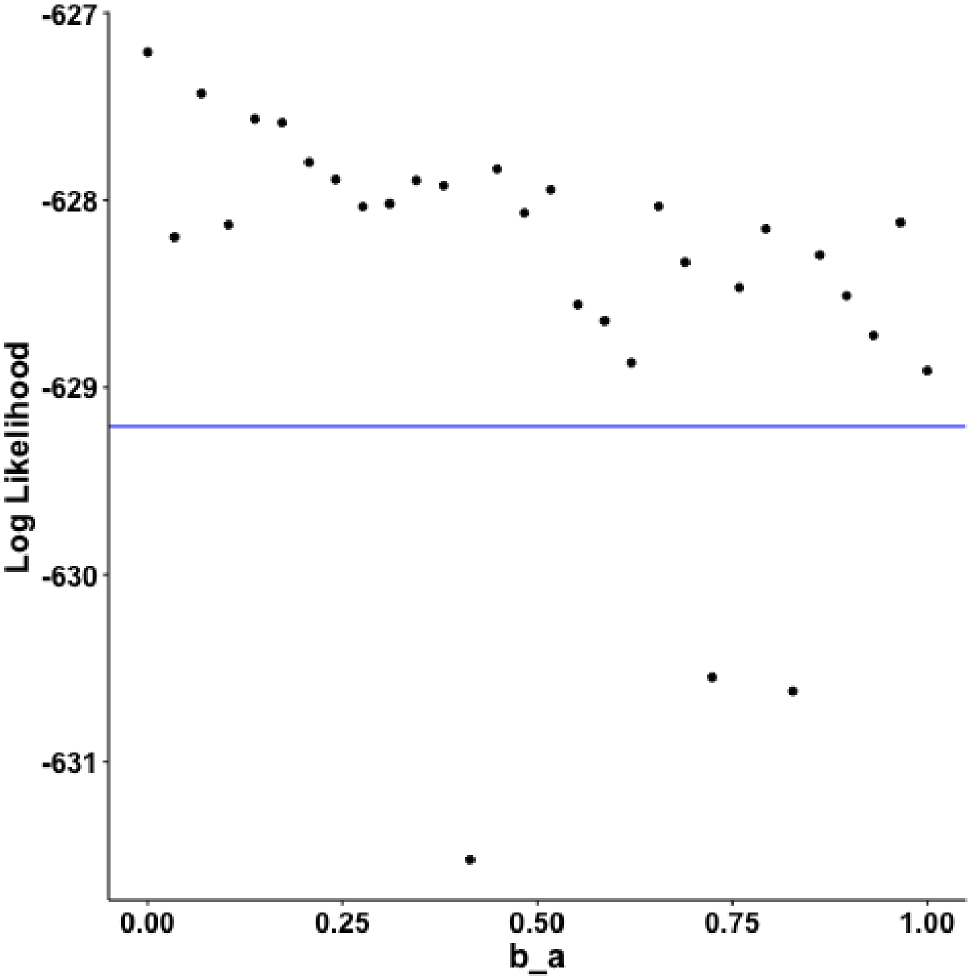

Figure 6: **Monte Carlo profile of the strength of transmission in asymptomatic cases relative to that of symptomatic cases ($b_{\mathrm{a}}$).** Each point represents the parameter combination from the Monte Carlo profile for $b_{\mathrm{a}}$ with the highest log-likelihood (with respect to observed cases) for a given value of $b_{\mathrm{a}}$. All points above the blue line are supported by the case data (i.e. they have likelihoods within 2 log likelihood units of the profile MLE).

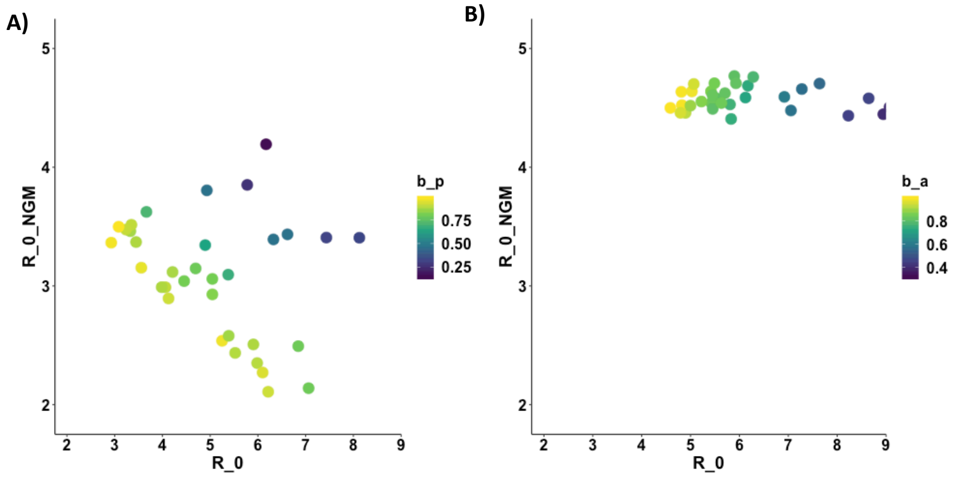

Figure 7: **Additional plots of the overall reproductive number ($R_{0_{\mathrm{NGM}}}$) vs the reproductive number in symptomatic individuals ($R_0$) from parameter combinations supported by case and serology data from the full SEPIAR (A) and SEIAR (B) models. A)** Each point represents one parameter combination within 2 log-likelihood units of the MLE (with respect to cases and serology) from the $b_{\mathrm{a}}$ profile using the full SEPIAR model. Each point is colored by the strength of pre-symptomatic transmission ($b_{\mathrm{p}}$). **B)** Each point represents one parameter combination within 2 log-likelihood units of the MLE (with respect to serology) from the grid search of the SEIAR model (no pre-symptomatic transmission). Points are colored by the relative strength of asymptomatic transmission ($b_{\mathrm{a}}$). For ease of plotting, in panel A) we exclude two parameter combination with very low pre-symptomatic transmission rates $b_p$. These outliers are also excluded from Figure 3 in the main manuscript , but are shown in Supporting Figure S8.

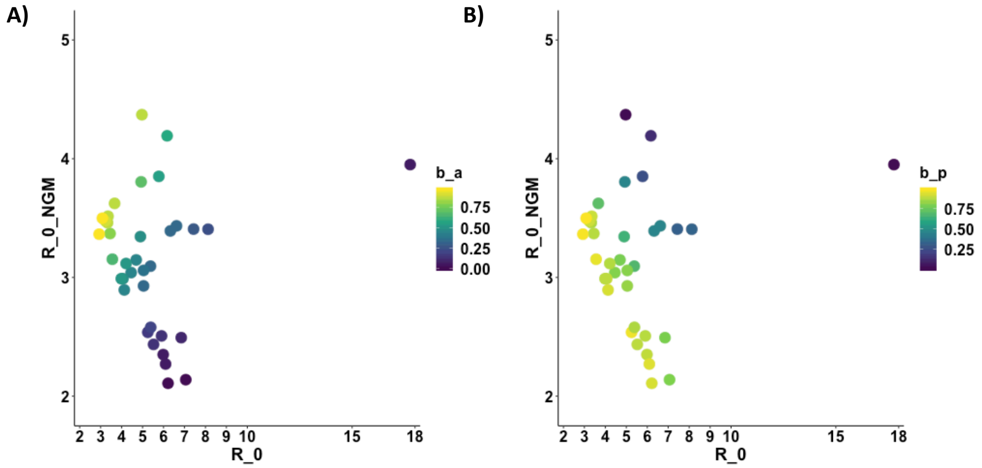

Figure 8: **Additional plots of the overall reproductive number ($R_{0_{\mathrm{NGM}}}$) vs the reproductive number in symptomatic individuals ($R_0$) including the outlier parameter combination colored by the relative strength of asymptomatic transmission ($b_{\mathrm{a}}$) (A) or the relative strength of pre-symptomatic transmission ($b_{\mathrm{p}}$) (B).** Each point represents one parameter combination within 2 log-likelihood units of the MLE (with respect to cases and serology) from the $b_{\mathrm{a}}$ profile of the full SEPIAR model. We include here the outlier parameter combinations with very high symptomatic $R_0$ greater than 15 that were excluded from Figure 3 and Figure S7.

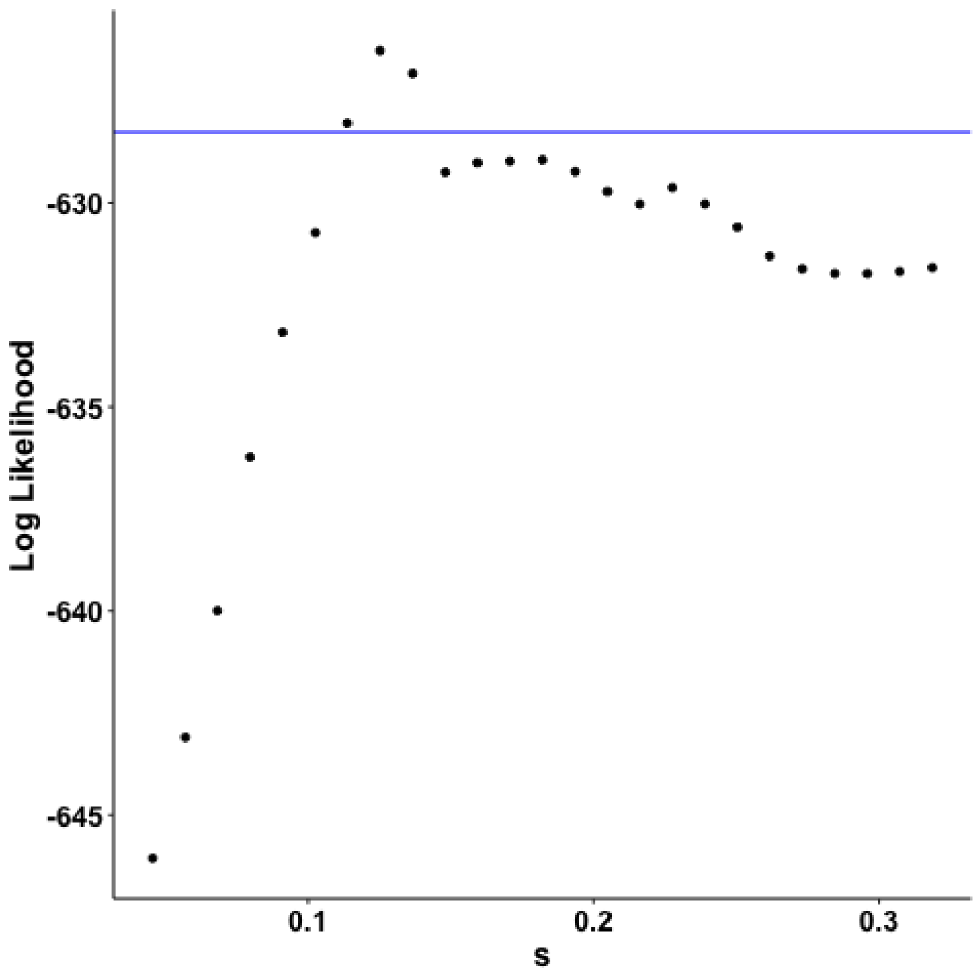

Figure 9: **Monte Carlo profile of the probability that an individual (who does not have COIVD-19) who shows up to the emergency department with respiratory symptoms is severe enough to merit testing for COVID-19 ($s$).** Each point represents the parameter combination from the Monte Carlo profile for the scaling parameter $s$ with the highest log-likelihood (with respect to observed cases) for a given value of $s$. All points above the blue line have likelihoods within 2-log likelihood units of the profile MLE).

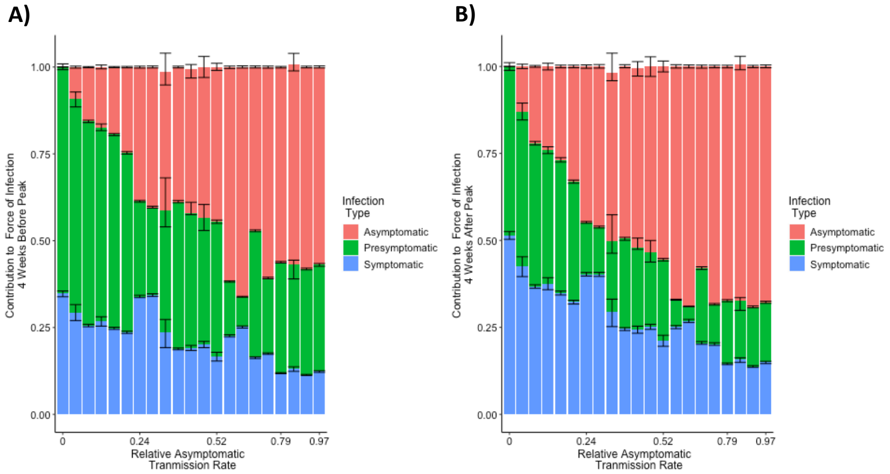

Figure 10: **Contributions from pre-symptomatic, symptomatic, and asymptomatic infections to the overall force of infection 4 weeks before (A) and after the peak in reported cases (B).** The calculation of the contribution to the overall force of infection from simulated trajectories of parameter combinations from the SEPIAR model supported by case and serology data as described in Figure 4 of the main manuscript was replicated using time points 4 weeks before and after the peak of reported cases on April 14, 2020 instead of at the time of the peak. As in Figure, two parameter combinations with rates of pre-symptomatic transmsison $b_p$ below 0.02 were excluded from the plot.

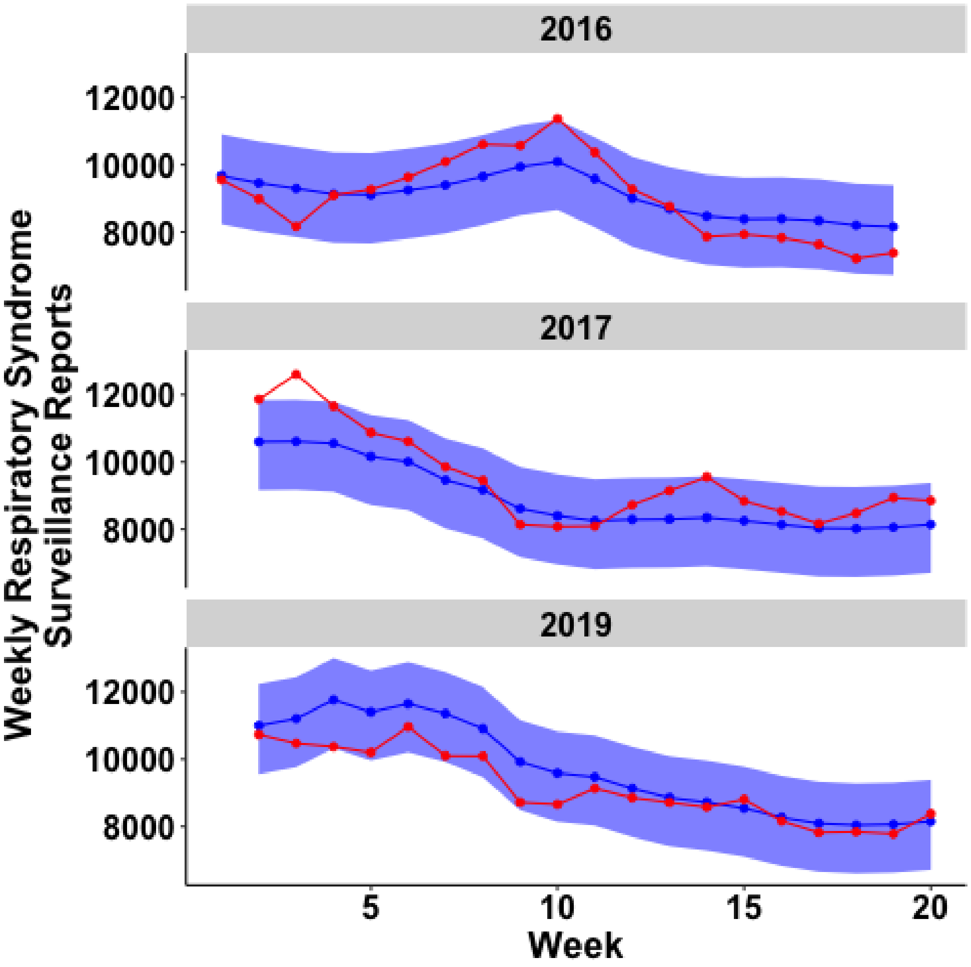

Figure 11: **Plot of observed respiratory syndrome surveillance reports compared to simulations from fitted statistical model** The red line corresponds to weekly respiratory infections from syndrome surveillance reports in NYC hospitals in 2016, 2017 and 2019 that were used to fit the statistical model in Section 4. The blue line represents the median estimate for the number of expected syndrome surveillance reports ($G(w, y)$ for that week and year from 100 simulations from the fitted statistical model. The shaded light blue region represents the 2.5% and 97.5% quantiles from those 100 simulations.

[6] Deparment of Health City of New York. Syndromic surveillance

data. 2020. URL https://a816-health.nyc.gov/hdi/epiquery/visualizations?PageType=ps&PopulationSource=Syndromic.

[7] New York State Department of Health. Influenza laboratory-confirmed cases by county: Beginning 2009-10 season. 2020. URL https://health.data.ny.gov/Health/Influenza-Laboratory-Confirmed-Cases-By-County-Beg/jr8b-6gh6.

[8] Centers for Disease Control and Prevention. Summary of the 2017-2018 influenza season. 2020. URL https://www.cdc.gov/flu/about/season/flu-season-2017-2018.htm.

[9] Centers for Disease Control and Prevention. Estimated influenza illnesses, medical visits, hospitalizations, and deaths in the united states — 2018–2019 influenza season. 2020. URL https://www.cdc.gov/flu/about/burden/2018-2019.html.
